# Disentangling the effects of nicotine versus non-nicotine constituents of tobacco smoke on major depressive disorder: A multivariable Mendelian randomization study

**DOI:** 10.1101/2024.06.25.24309292

**Authors:** Chloe Burke, Gemma Taylor, Tom P Freeman, Hannah Sallis, Robyn E Wootton, Marcus R Munafò, Christina Dardani, Jasmine Khouja

## Abstract

**Background and aims:** There is growing evidence that tobacco smoking causes depression, but it is unclear which constituents of tobacco smoke (e.g., nicotine, carbon monoxide) may be responsible. We used Mendelian randomisation (MR) to examine the independent effect of nicotine on depression, by adjusting the effect of nicotine exposure (via nicotine metabolite ratio [NMR]) for the overall effect of smoking heaviness (via cigarettes per day [CPD]) to account for the non-nicotine constituents of tobacco smoke.

**Design:** Univariable MR and multivariable MR (MVMR) were used to explore the total and independent effects of genetic liability to NMR and CPD on major depressive disorder (MDD). Our primary method was inverse variance weighted (IVW) regression, with other methods as sensitivity analyses.

**Setting and participants:** For the exposures, we used genome-wide association study (GWAS) summary statistics among European ancestry individuals for CPD (n=143,210) and NMR (n=5,185). For the outcome, a GWAS of MDD stratified by smoking status was conducted using individual-level data from UK Biobank (n=35,871-194,881).

**Measurements:** Genetic variants robustly associated with NMR and CPD.

**Findings:** Univariable MR indicated a causal effect of CPD on MDD (odds ratio [OR]^IVW^=1.13, 95% confidence interval [CI]=1.04-1.23, *P*=0.003) but no clear evidence for an effect of NMR on MDD (OR^IVW^=0.98, 95% CI=0.97-1.00, *P*=0.134). MVMR indicated a causal effect of CPD on MDD when accounting for NMR (OR^MVMR-IVW=^1.19, 95% CI=1.03-1.37, *P*=0.017; OR^MVMR-EGGER=^1.13, 95% CI=0.89-1.43, *P*=0.300) and weak evidence of a small effect of NMR on MDD when accounting for CPD (OR^MVMR-IVW=^0.98, 95% CI=0.96-1.00, *P*=0.056; OR^MVMR-EGGER=^0.98, 95% CI=0.96-1.00, *P*=0.038).

**Conclusions:** The causal effect of tobacco smoking on depression appears to be largely independent of nicotine exposure, which implies the role of alternative causal pathways.

## INTRODUCTION

Tobacco smoking is amongst the leading causes of preventable disease and death worldwide [1]. An estimated 1.14 billion adults globally smoke tobacco products regularly [2]. Although tobacco smoking in high income countries (e.g., US, UK) has markedly decreased in recent decades [3,4], smoking prevalence amongst individuals with mental health conditions remains approximately twice as high [5]. Major Depressive Disorder (MDD) is a highly prevalent psychiatric condition with an estimated lifetime prevalence of ∼14.6% for adults in high-income countries [6] and is a leading cause of global disease burden [7,8]. However, knowledge of actionable preventative strategies that could mitigate depression risk remains limited [9].

Considerable debate has surrounded the association between smoking and mental illness [10], with several proposed explanations: (i) smoking may causally impact mental health; (ii) those with poorer mental health may use tobacco/nicotine to self-medicate (i.e., the self-medication model); (iii) smoking and mental health may share one or more common risk factors (e.g., genetic, environmental) [11]. These explanations are not mutually exclusive, and each has distinct key implications for policy and practice. There is growing evidence across multiple study designs (e.g., longitudinal observational analyses, Mendelian randomisation [MR], smoking cessation interventions) that tobacco smoking causes worse mental health [12–15], including depression [16,17]. However, it remains unclear which constituents of tobacco smoke (e.g., nicotine, carbon monoxide) may confer negative effects for mental health [18].

Multiple underlying biological mechanisms for a potential causal relationship between tobacco smoking and depression have been proposed [10]. A key theory relates to neuroadaptations in nicotinic pathways in the brain, whereby stimulation of nicotine receptors augments the release of neurotransmitters (e.g., dopamine, serotonin, norepinephrine) which have been implicated in the aetiology of depression [19,20]. In support of the potential role of nicotine exposure in mental illness, there is evidence from observational studies that e-cigarette (i.e., electronic devices which often contain nicotine) use is associated with increased risk of mood disorders [21–23]. Furthermore, the reported benefits of smoking cessation for mental health [15] appear to be reduced in former smokers that report recent (i.e., past month) e-cigarette use [24,25]. However, due to the inherent limitations of observational methods (e.g., reverse causation, residual confounding) inferring causality from these studies is difficult. Furthermore, few never smokers regularly use e-cigarettes [26–28], thus confounding by exposure to tobacco smoke and its other constituents is likely.

Randomised controlled trials (RCTs) are the gold-standard for exploring causal relationships [29], however an RCT of long-term nicotine exposure in non-smokers would be both unethical and impractical. MR is a useful, alternative approach to studying causal relationships where an RCT is unfeasible [30]. MR employs common genetic variants as instrumental variables to explore causal relationships between exposures and health outcomes [31]. Provided several key assumptions are met [32,33], MR can minimise bias due to reverse causation and residual confounding as the genetic variants used as proxies for the exposure of interest are randomly assigned at meiosis and fixed at conception [30].

MR has previously been employed using genetic variants robustly associated with tobacco smoking phenotypes (e.g., smoking initiation, lifetime smoking index, smoking heaviness) to estimate the total causal effect of a smoking-related exposure on various mental health outcomes [14]. However, as there are no published large genome-wide association studies (GWAS) of nicotine exposure without exposure to tobacco smoking (e.g., e-cigarette use amongst non-smokers), disentangling the potential causal effects of nicotine from the non-nicotine constituents of tobacco smoking has been limited due to the potential shared genetic aetiology of tobacco smoking and e-cigarette use behaviours [34].

A novel framework, employing multivariable MR (MVMR), has recently been applied to explore the effects of nicotine and non-nicotine constituents of tobacco smoking for various physical health outcomes (e.g., lung cancer, chronic obstructive pulmonary disease) [35]. MVMR is an extension to MR that includes multiple exposures to estimate the effect of one exposure (e.g., smoking heaviness), independent of other, genetically correlated, exposures (e.g., nicotine metabolite ratio) [36–38]. MVMR is therefore a valuable tool to explore highly correlated phenotypes, such as tobacco smoke exposure and nicotine intake. GWAS have previously identified genetic variants associated with smoking heaviness (i.e., cigarettes per day) [39] and the nicotine metabolite ratio (NMR] [40] – the ratio of 3’hydroxycotinine (3HC)/cotinine (COT) [41].

NMR is a biomarker which represents the rate of nicotine metabolism [42]; smokers with a higher NMR will clear nicotine more quickly from their system. However, having a higher NMR is associated with smoking more cigarettes per day (CPD) (Figure 1) [43]. This means that the effect of NMR in a univariable MR analysis would be ambiguous.

**Figure 1.**
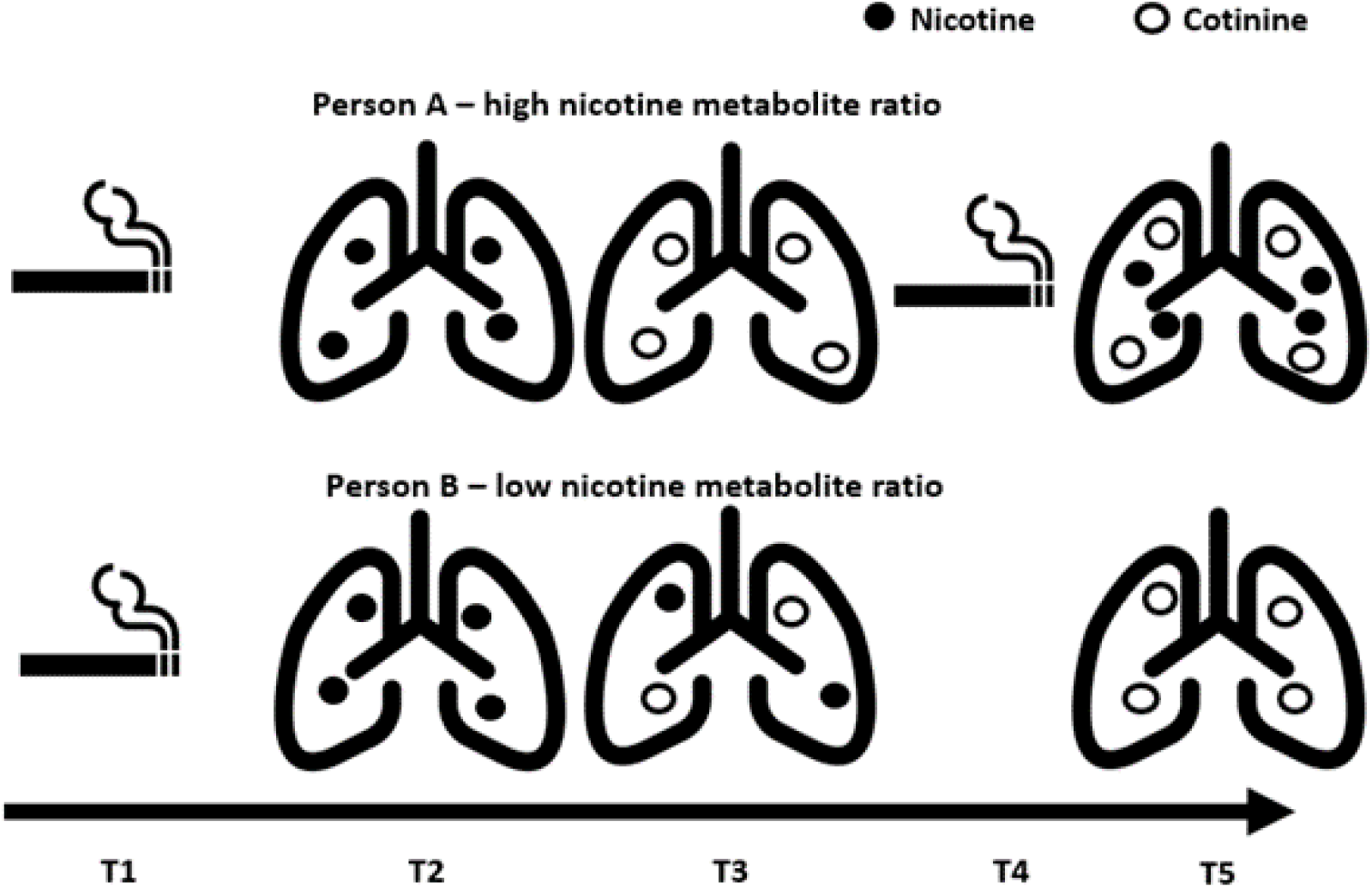
Illustration of the impact of nicotine metabolite ratio on circulating nicotine. *Note.* This image is reused, with permission, from authors from Khouja and colleagues [35]. Illustration depicts the differences in circulating nicotine between two people who smoke, with a different NMR. Person A has a high NMR and Person B has a low NMR. At timepoint 1 (T1) both individuals smoke one cigarette inhaling the same amount of nicotine, as shown at timepoint 2 (T2). Subsequently, at timepoint 3 [T3], Person A will have less circulating nicotine in their body than Person B given the same nicotine exposure as more nicotine has been metabolised into cotinine. Because smokers with a higher NMR clear nicotine more quickly from their system, this will often result in them smoking more cigarettes per day at timepoint 4 (T4), whereas a smoker with a lower NMR may not. This results in Person A having more circulating nicotine in their body than Person B over the same time period. This figure includes images from the following sources in accordance with their copyright licence: https://commons.wikimedia.org/wiki/File:Bootstrap_lungs.svg; https://commons.wikimedia.org/wiki/File:Noun_146.svg. The lung images have been adapted to include circles. https://doi.org/10.1371/journal.pgen.1011157.g001

By including both NMR and CPD in an MVMR model (Figure 2), the effect of nicotine exposure per cigarette smoked is fixed and the effect of nicotine exposure per cigarette can be explored [35]. For a full explanation of the model, please refer to Khouja and colleagues [35].

**Figure 2.**
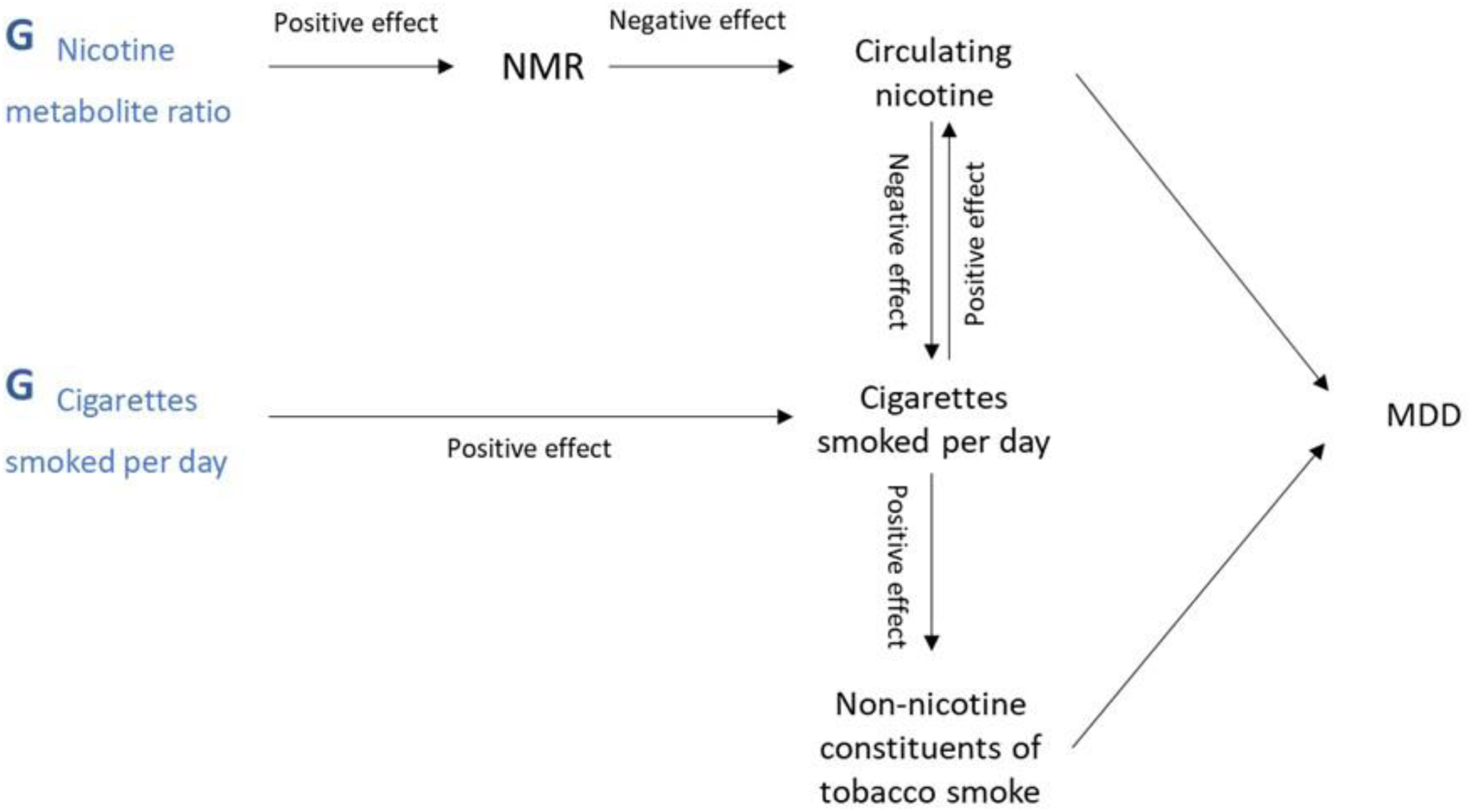
Schematic of the study model. *Note.* Schematic adapted from Khouja and colleagues [35], with permission, to reflect outcome of interest. NMR=Nicotine Metabolite Ratio; MDD=Major Depression. G=genetic variants associated with the named exposure. Where causal directions between variables are known (i.e., evidenced in previous research), the direction of the effect is indicated as ‘positive’ or ‘negative’. A higher genetic liability (G-NMR, G-CPD [cigarettes per day]) has a positive, or increasing, effect on NMR and CPD (i.e., higher NMR and greater number of CPD). A higher NMR has a negative, or decreasing, effect on circulating nicotine as an individual with a higher NMR has a faster rate of nicotine metabolism.

**Figure 3.**
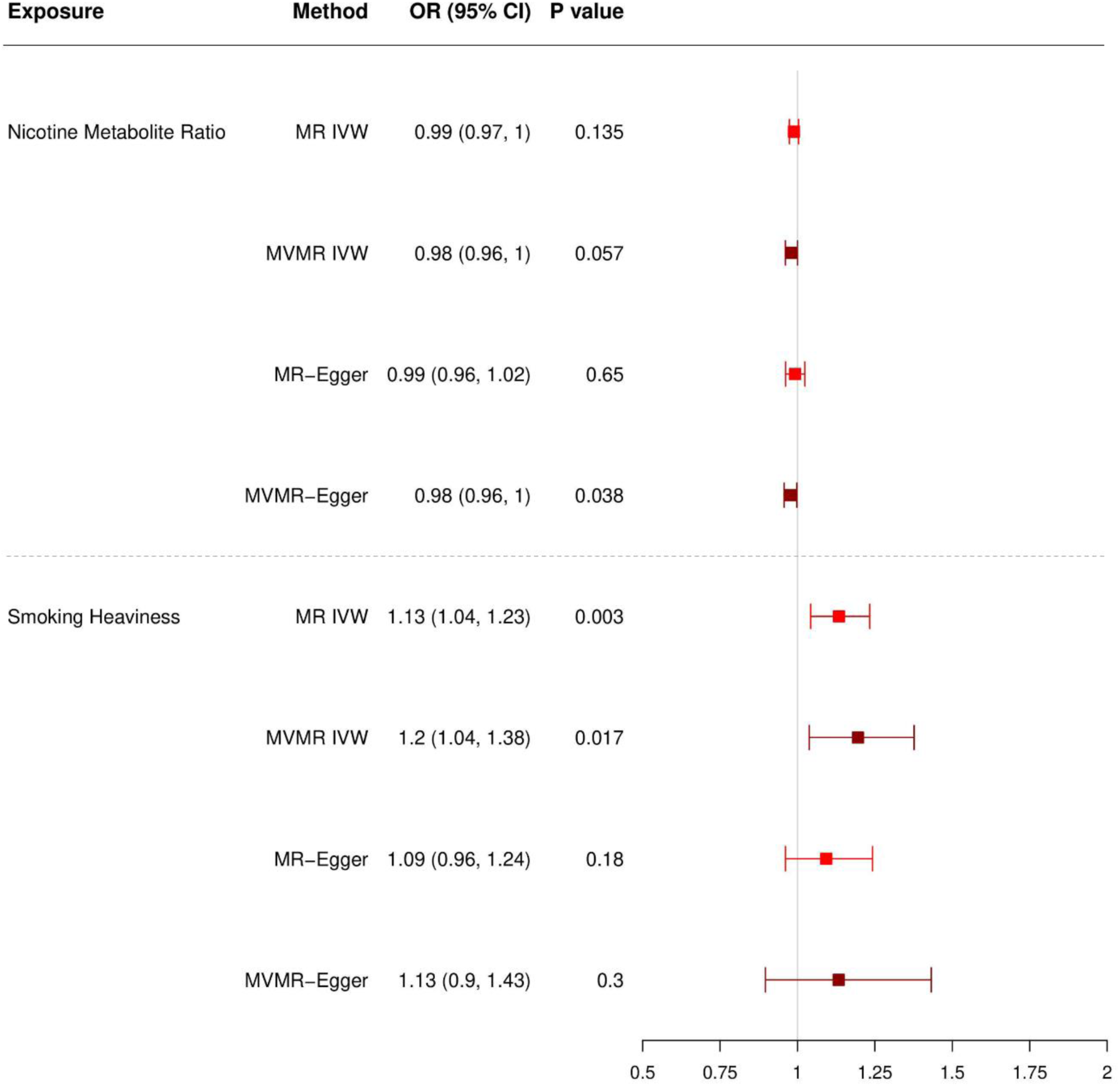
Forest plot displaying the effect of NMR and smoking heaviness on MDD: univariable and multivariable MR (MVMR) results among ever smokers. *Note.* OR=Odds Ratio. CI=Confidence Interval. MR=Mendelian Randomisation. MVMR=Multivariable MR. IVW=Inverse Variance Weighted. Light red = univariable MR. Dark red = multivariable MR.

Identifying which constituents of tobacco smoking are causally related to mental illness has been identified as a priority for future research [10,18], and has the potential to inform current and future perspectives on tobacco smoking policies. We employed a summary-level (i.e. two-sample) MVMR framework to explore the independent causal effects of nicotine compared to the non-nicotine constituents of tobacco smoke on MDD.

## METHODS

Our statistical analysis plan was pre-registered (https://osf.io/udqkm; 18/12/2023). See supplementary materials for protocol deviations (Supplementary Note S1).

### Ethics statement

UK Biobank received ethics approval from the North West Multi-Centre Research Ethics Committee as a Research Tissue Bank approval (REC; 11/NW/0382). Approval to use these data was sought and approved by UK Biobank (Project ID: 9142).

### Data sources

Summary-level genetic data from well-powered, published GWAS were used to identify relevant instrumental variables (IVs) for NMR and smoking heaviness (measured via CPD) [39,40]. Both GWAS were performed amongst smokers; the NMR GWAS was performed among current smokers [40] and the CPD GWAS was performed among ever smokers (i.e., current and former smokers) [39]. Information regarding the contributing cohorts, periods of recruitment, genotyping, imputation, and quality control are described in detail in the original meta-analyses [39, 40]. To the best of our knowledge, there were not existing available GWAS for depression that were appropriately stratified (i.e., stratified by ever versus never smoking status), which is necessary to meet the assumption that summary-level MR samples are drawn from the same underlying population [30,44]. As such, we used UK Biobank data to conduct a GWAS of major depressive disorder (MDD) which was stratified by smoking status and restricted to individuals of European ancestry. GWAS were stratified by whether participants had (1) ever smoked, and further stratified into (2) current smoker, or (3) former smoker, with (4) never smoker as the comparator. GWAS were performed using the MRC Integrative Epidemiology Unit UK Biobank GWAS pipeline (V2) [45,46].

#### Smoking heaviness

Liu and colleagues [39] report summary-level statistics from a GWAS meta-analysis of smoking heaviness (measured by standard deviation [SD] change in CPD categories, equivalent to 2-3 additional CPD) among 337,334 ever smokers of European ancestry. To eliminate sample overlap with our GWAS of MDD, summary statistics were obtained with UK Biobank (n=120,744) removed. 23&Me participants (n=73,380) were also excluded from the summary data due to data sharing restrictions. The remaining sample consisted of data from 143,210 participants. Smoking heaviness was defined as the average number of cigarettes smoked per day (e.g., *“How many cigarettes do/did you smoke per day?”*) as a current or former smoker. Self-reported quantities were binned (1-5 CPD, 6-15 CPD, 16-25 CPD, 26-35 CPD, 36+ CPD) or else pre-defined contributing cohort bins were used [39]. Analyses were adjusted for age, sex, genetic principal components and smoking status (i.e., current versus former). SNPs were reported as independent if they explained additional variance in conditional analyses using a partial correlation-based score statistic [47]. The 55 genome-wide significant conditionally independent SNPs associated with CPD explained ∼4% of the variance in CPD.

#### Nicotine metabolite ratio

Buchwald and colleagues [40] report summary-level statistics from a GWAS meta-analysis of NMR (measured by SD change in NMR), among 5,185 current smokers with cotinine levels ≥10ng/ml (i.e., indicative of recent smoking) of European descent. NMR is a ratio of 3’hydroxycotinine/cotinine and indicates how quickly a person metabolises and clears nicotine [41]. Analyses were adjusted for population substructure, age, sex, BMI, alcohol use and birth year. SNPs were reported as independent if they explained additional variance in a stepwise conditional regression using genome-wide complex trait analysis (GCTA) [48]. The seven genome-wide significant conditionally independent SNPs associated with NMR explained ∼38% of the variance in NMR.

#### Major depressive disorder

We conducted a GWAS of MDD using data from UK Biobank, a population-based cohort consisting of ∼500,000 people aged between 37 and 73 years recruited between 2006 and 2010 from across the UK [49]. Participants provided extensive information about their lifestyle, physical measures and had blood, urine and saliva samples collected and stored for future analysis. Participants attended a baseline assessment, and subsets of participants completed repeat assessments including an online mental health questionnaire (MHQ) in 2017 [50]. A detailed description of the study design, participants and quality control (QC) methods has been reported previously [49–51].

The full data release contains the cohort of successfully genotyped samples (n=488,377). Analyses were restricted to individuals of ‘European’ ancestry as defined by an in-house k-means cluster analysis [45]. Information about genotyping and imputation can be found in Supplementary Note S2. The GWAS were conducted using the linear mixed model (LMM) association method as implemented in BOLT-LMM (v2.3) [52] and were adjusted for age, sex and genotype array [45]. Betas and their corresponding standard errors were transformed to log odds ratios and their corresponding 95% confidence intervals [52]. Smoking status was categorised using self-reported information on smoking status (ID: 20116). Using this variable an ‘ever smokers’ category was derived, defined as currently or previously smoking occasionally, most days or daily (i.e., more than once or twice). Current smoking was defined as currently smoking occasionally, most days or daily. Former smoking was defined as not currently smoking, but previously smoking occasionally, most days or daily. Individuals who reported trying smoking once or twice, or reported never smoking, were categorised as never smokers.

MDD cases were identified following a validated approach to defining lifetime major depression in UK Biobank [53]. Full details, and UK Biobank field IDs, are provided in Supplementary Notes S3–S5. Briefly, ‘non-MHQ’ cases were identified if endorsing at least two measures of depression from: ‘help-seeking’, ‘self-reported depression’, ‘antidepressant usage’, ‘depression (Smith)’ [54] or ‘hospital (ICD-10) depression’. ‘MHQ cases’ were identified from the online follow-up if they met criteria for lifetime MDD, as assessed by the Composite International Diagnostic Interview–Short Form (CIDI-SF) which approximates DSM criteria [50]. Controls comprised individuals who did not endorse any of the depression phenotypes (i.e., excluding those endorsing one case indicator at baseline) or any of five psychosis phenotypes: ‘self-reported psychosis’, ‘antipsychotic usage’, ‘bipolar (Smith)’ [54], ‘hospital (ICD-10) psychosis’ and ‘psychosis (MHQ)’. Flowcharts of samples contributing to each stratified GWAS are provided in Supplementary Figures S1–S4.

To explore population stratification, SNP-based heritabilities (Supplementary Table S1) were calculated using linkage-disequilibrium score regression (LDSC v1.0.1) [55, 56] and QQ plots were generated (Supplementary Figure S5).

Statistical analysis

All analyses were conducted using R version 4.3.1. Statistical analysis was completed using the *TwoSampleMR*, *MVMR* and *MendelianRandomization* packages [57–59] and the statistical code and data used is available online (https://doi.org/10.5281/zenodo.12187311).

#### Selection of genetic variants

SNPs that were identified as conditionally independent at the genome-wide significant level (*p* < 5 × 10^-8^) in the GWAS analyses of NMR and CPD were selected for inclusion in the analysis [39,40], which yielded 55 SNPs associated with smoking heaviness and 7 SNPs associated with NMR. All identified SNPs were available in the outcome GWAS of MDD. After harmonising, the combined exposure datasets (i.e., SNPs for CPD and NMR) were clumped (LD R^2^ <0.1, >500kb) to remove overlapping loci and ensure overall independence for the MVMR analysis. Supplementary Table S2 details which SNPs were included and provides reasons for exclusion. Supplementary Note S6 provides additional detail on the harmonisation and clumping methods.

#### Univariable MR

To provide a comparison for the MVMR analysis, the total effect of NMR and smoking heaviness on MDD were examined using univariable MR. The main method was inverse variance weighted (IVW) regression [60]. We investigated how robust estimates were to violations of the exclusion restriction instrumental variable (IV) assumption (i.e., no horizontal pleiotropy) with MR-Egger [61], weighted median [62] and weighted mode [63] applied as sensitivity methods. Consistent results (i.e., direction, size of effects) across these estimators suggest findings are less likely to be driven by pleiotropic or heterogeneous effects. Details on the assumptions of MR and the univariable analysis methods are reported in Supplementary Note S6.

We computed Cochran’s Q to assess heterogeneity between SNP-estimates in each instrument and the F-statistic to assess instrument strength, whereby F >10 suggests lower risk of weak instrument bias and Q-statistic should be less than the number of SNPs [64]. To assess whether the NO Measurement Error (NOME) assumption (defined in Supplemental Note 6) was satisfied for MR-Egger, we computed the I^2^_Gx_ statistic, with values >0.9 indicating low risk of bias due to measurement error [65]. We additionally generated scatter plots of SNP effect sizes, and performed leave-one-out analyses to further explore heterogeneity and potential pleiotropic effects [33]. Steiger filtering, which computes the amount of variance each SNP explains in the exposure and outcome variables, was conducted to confirm the direction of effect [66] (i.e., identify SNPs which are more predictive of the outcome than the exposure).

#### Multivariable MR

The independent effects of NMR and smoking heaviness on MDD were explored using two complementary MVMR methods: MVMR-IVW and MVMR-Egger [38,67]. For MVMR-Egger, results are reported for analyses with SNPs oriented with respect to each exposure of interest [68].

An additional assumption for MVMR is that instruments are strongly associated with each exposure given the other exposures included in the model, or ‘conditional relevance’ [38]. In univariable MR, weak instruments will bias estimated effects in the direction of the observational estimate; however, in MVMR, it is unclear which direction of bias will occur due to weak instruments [38]. The conditional F-statistic (F_TS_) for summary-level MR was calculated to evaluate the strength of the SNP-exposure associations, conditional on other exposures (i.e., whether SNPs jointly predict NMR after predicting smoking heaviness, and vice versa), with values ≥10 indicating results are unlikely to suffer from weak instrument bias [58]. A modified form of Cochran’s *Q* statistic for MVMR was applied to detect heterogeneity among the SNPs included, where *Q* estimates should be less than the number of SNPs included in the model to indicate no excessive heterogeneity [58].

For both univariable and MVMR, analyses were performed amongst never smokers as a negative control analysis to explore potential bias from horizontal pleiotropy (i.e., effects observed among never smokers could indicate SNPs influencing the outcome directly or via another phenotype, but not via the target exposure).

## RESULTS

### Descriptive statistics

In GSCAN, information on cigarettes smoked per day, either current or former average, was collected from 337,334 current and former smokers with an average binned level of 16-25 CPD [39]. In the study by Buchwald and colleagues [40], NMR data were collected from 5,185 current smokers with an average of 11.5 (SD 6.0) to 20.3 (SD 7.9) CPD and average NMR of 0.41 (SD 0.22) to 0.47 (SD 0.24) across the five contributing cohorts. The average case prevalence of MDD amongst the stratified smoking groups in UK Biobank was: 25.5% (ever smokers), 30.7% (current smokers), 24.0% (former smokers) and 20.3% (never smokers).

### Instrument strength and heterogeneity

The conditional F-statistics indicated that the SNPs used in these analyses are strong (F > 10) instruments for assessing the independent effects of smoking heaviness while accounting for the effect of NMR (F=23.13) and for assessing the independent effects of NMR while accounting for smoking heaviness (F=26.16; Supplementary Table S3).

Cochran’s Q statistics (Supplementary Table S3) and scatter plots (Supplementary Figures S6–S7) indicated heterogeneity for some analyses, highlighted in each relevant results section. As such, both univariable and multivariable analyses are interpreted alongside methods to explore pleiotropy (i.e., MR-Egger and MVMR-Egger) which generate estimates robust to directional horizontal pleiotropy under the assumption that the exposure effects of the individual SNPs are independent of their pleiotropic effects on the outcome (i.e., InSIDE assumption) [69]. However, MR-Egger and MVMR-Egger methods have much lower statistical power when compared to other methods. Therefore, we focus on consistency in the direction, rather than supporting statistical evidence for the effect.

### Univariable MR

Results are presented as odds ratios (OR) per standard deviation (SD) increase in the exposure phenotype (i.e., per SD increase in the NMR or cigarettes per day). ORs are presented for inverse variance weighted (OR^IVW^), MR-Egger (OR^EGG^), weighted median (OR^MED^) and weighted mode (OR^MOD^) analyses.

Among ever smokers, the MR-IVW results provided no clear evidence that NMR affects MDD risk (OR^IVW^=0.99, 95% CI 0.97–1.00; *p=*0.135; Supplementary Table S3). This was supported by the MR-Egger, weighted median and weighted mode results (OR^EGG^=0.99, 95% CI 0.96–1.02, *p=*0.650; OR^MED^=0.99, 95% CI 0.97–1.01, *p=*0.273; OR^MOD^=0.99, 95% CI 0.97–1.01, *p=*0.297; respectively; Supplementary Table S4).

The MR-IVW results provided evidence that increased smoking heaviness increases MDD risk (OR^IVW^=1.13, 95% CI 1.04–1.23, *p=*0.003; Supplementary Table S3), and MR-Egger, weighted median and weighted mode showed consistent effects (OR^EGG^=1.09, 95% CI 0.96– 1.24, *p=*0.180; OR^MED^=1.12, 95% CI 1.02–1.23, *p=*0.018; OR^MOD^=1.13, 95% CI 1.04–1.22, *p=*0.004; respectively; Supplementary Table S4).

Although there was evidence of considerable heterogeneity in the smoking heaviness analyses indicated by the Q-statistic (*Q*=80.62; Supplementary Table S3) and scatter plot (Supplementary Figure S7), direction of effect was supported by the MR-Egger results and the intercept indicated heterogeneity was likely not due to directional horizontal pleiotropy (MR-Egger intercept *p=*0.456; Supplementary Table S3). Additionally, there was no strong evidence of pleiotropy or bias due to population stratification indicated by the results among never smokers (Supplementary Table S3; Supplementary Figure S8) and leave-one-out analyses suggested overall effect estimates were not driven by one particular SNP (Supplementary Figures S11–S12).

### Multivariable MR

Among ever smokers, the MVMR-IVW results provided weak evidence of a small causal effect of NMR on MDD risk (OR^IVW^=0.98, 95% CI 0.96–1.00, *p=*0.057; OR^EGG^=0.98, 95% CI 0.96–1.00, *p=*0.038; Supplementary Table S3). The MVMR-IVW results provided evidence that increased smoking heaviness increases MDD risk (OR^IVW^=1.20, 95% CI 1.04–1.38, *p=*0.017) and was supported by the direction of effect in the MVMR-Egger analyses (OR^EGG^=1.13, 95% CI 0.90–1.43, *p=*0.300; Supplementary Table S3).

There was evidence of heterogeneity indicated by the Q-statistic (*Q*=68.49; Supplementary Table S3), but direction of effect was supported by the MR-Egger results, the intercepts indicated heterogeneity was likely not due to directional horizontal pleiotropy (MVMR-Egger intercepts *p=*0.368–0.812) and there was no clear evidence of an effect among never smokers (Supplementary Table S3; Supplementary Figure S8).

### Supplementary and sensitivity analyses

Among ever and never smokers, Steiger filtering indicated no clear evidence to support reverse causality (i.e., depression influencing smoking heaviness). All the genetic instruments for CPD and NMR explained more variance in the exposure instrument than in MDD (Supplementary Table S5).

The results among current and former smokers can be found in Supplementary Tables S3-S4. Analyses across both groups indicated no clear evidence for an independent or total casual effect of NMR on MDD, and slightly weaker evidence for an independent and total causal effect of CPD on MDD amongst current smokers (Supplementary Note S7; Supplementary Figures S9-S10).

## DISCUSSION

In this study, we employed a summary-level MR approach to test putative causal effects of nicotine exposure and smoking heaviness on MDD. We used univariable MR and multivariable MR to distinguish the independent effects of nicotine from the independent effects of non-nicotine components of tobacco smoke, employing genetic variants robustly associated with NMR and CPD as proxies, as has been applied in previous work to examine the effects of smoking on various physical health outcomes (e.g., lung cancer) [35].

Our results suggest no clear evidence for a total causal effect of nicotine exposure on MDD, although MVMR analysis provided weak evidence for a small, potentially negligible, independent effect of NMR on MDD, such that increased genetic liability to NMR (i.e., decreased nicotine exposure per cigarette smoked) reduced the risk of MDD. We found evidence consistent with a total causal effect of increased smoking heaviness on MDD and after adjusting for NMR, the magnitude of effect was slightly larger, although with overlapping confidence intervals. Together these results suggest we cannot rule out nicotine as a potential mechanism underlying the effect of tobacco smoking on depression. However, the observed effects of smoking heaviness after adjusting for NMR indicate the involvement of other causal pathways beyond nicotine exposure. Results obtained were consistent across different sensitivity analyses performed that rely on different assumptions, providing stronger evidence to support the potential causal role of tobacco smoking on MDD [70].

If the observed causal effect of tobacco smoking on MDD was largely independent of nicotine exposure there are several mechanisms, biological and non-biological, which could explain the effect [10, 71]. A plausible biological mechanism relates to inflammation and oxidative stress. Uptake of toxins in cigarette smoke (e.g., fine particulate matter, heavy/transition metals) can lead to neuroinflammation and cerebral oxidative stress [10, 71]. Smoking behaviour is associated observationally with both depression and increased oxidative stress biomarkers [72, 73] and evidence from a multivariable MR analysis of lifetime smoking, depression and inflammation, proxied via IL-6 activity, suggests that the causal effect of smoking on depression is substantially attenuated when accounting for the effects of inflammation [74]. Alternatively, a plausible non-biological mechanism relates to the effect of tobacco smoking on MDD via social pathways such as social isolation or loneliness. Smoking has been found to increase social isolation [75, 76] and loneliness [75, 77]. There are various reasons why smoking may increase social isolation and loneliness (e.g., development of smoking-related illness, perceived stigma, legislative restrictions on smoking in public places, effect of smoking on mobility and functional capacity) [75], which are, in turn, associated with increased risk of MDD [77, 78].

Another potential explanation for the difference in results between smoking heaviness and NMR, which doesn’t relate to differences in underlying true causal effects, is horizontal pleiotropy in the smoking heaviness instrument which is a particularly important issue in MR studies of behavioural risk factors and psychiatric outcomes [79]. Although evidence for horizontal pleiotropy has been found to a lesser extent for smoking heaviness versus smoking initiation [80], caution is still advised when using this instrument and results should be interpreted alongside pleiotropy-robust sensitivity methods (e.g., MR-Egger) and alternative approaches (e.g., negative control analyses) as we have applied in this study. Results for smoking heaviness were consistent across pleiotropy robust methods, with no strong evidence for directional horizontal pleiotropy indicated by the MR-Egger intercept or analyses amongst non-smokers.

Importantly, smoking heaviness is a time-varying exposure and genetic influences for smoking behaviour are not static over age [81]. In this scenario, MR estimates can be interpreted as the lifetime effect of being on a trajectory for the exposure associated with having an exposure level that is a unit higher at the time it is measured [82]. While GWAS procedures for identifying smoking heaviness SNPs control for age [39,81], this does not account for potential developmental sensitivity [81]. For example, it has been demonstrated that the genetic risk for CPD relates to adult, but not adolescent smoking heaviness [83]. Furthermore, the incidence of many mental health conditions is not uniformly distributed across the lifespan. For example, MDD is typically developed in early adulthood, with an estimated median age of onset of 31 years [84]. As such, our results will not capture any time-sensitive effects (e.g., exposure to higher levels of nicotine during critical development periods such as adolescence).

### Strengths and limitations

Our study has several strengths, including: (i) the application of a genetically informed approach to strengthen causal inference; (ii) the use of a smoking heaviness phenotype to examine the effect of frequency of use on risk of depression; (iii) the use of several sensitivity analyses and robust MR methods to examine the validity of core MR assumptions; and (iv) the inclusion of a negative control exposure.

However, the results should be interpreted considering some important limitations. First, a limitation which applies to all MR studies is that results may be impacted by selection bias. This may occur if both smoking heaviness and MDD influence selection into our study. The participants of UK Biobank are substantially better educated, healthier (e.g., fewer chronic health conditions) and less likely to smoke than the general population [85], which can distort genetic associations and downstream analyses (e.g., MR estimates) especially for socio-behavioural traits such as smoking [86]. Future GWAS using UK Biobank should consider the application of inverse probability weighting to correct for participation bias in this cohort [86]. Second, genetic variants linked to smoking heaviness are correlated with cognitive (e.g., years of education) [39,87] and physical traits (e.g., obesity) [39] which are associated with mental health and could be possible sources of pleiotropy. Whilst we employed methods to explore horizontal pleiotropy, future research could further explore the role of pleiotropic effects using MVMR to test the independent effects of tobacco smoking controlling for potential confounding factors, or novel PheWAS-based clustering of MR instruments [88]. Third, although multi-ancestry GWAS of smoking heaviness and major depression are available [89,90] the NMR GWAS was restricted to individuals of European ancestry and the MDD GWAS would not be stratified by smoking status, leading to violations of an assumption of MVMR is that all data included in the model are drawn from the same underlying population [30,44]. Thus, our findings may not be generalizable to other populations. Finally, although MR represents a useful methodology to explore causal effects, results from MR studies should not be interpreted in isolation and should be triangulated with results from studies using different and unrelated key sources of bias [91]. One potential avenue of research could be to conduct secondary analyses of data from e-cigarette cessation trials, where changes in depressive symptoms are compared between cessation and continued use, as has been applied to smoking cessation trials [15]. Alternatively, secondary analyses of data from trials of reduced nicotine content cigarettes could be used to compare depressive symptoms between participants using lower versus higher nicotine content cigarettes.

## CONCLUSION

Our study provides supporting evidence of a causal effect of tobacco smoking on depression, as reported in previous MR studies using different exposure instruments (e.g., lifetime smoking, smoking initiation). To some extent, our results suggest that the causal effect of tobacco smoking on depression is largely independent of nicotine exposure and implies the role of alternative causal pathways (e.g., driven by other constituents of tobacco smoke, non-biological pathways). Although further research is required to triangulate these findings [91], our results underpin the importance of strategies to prevent individuals from initiating tobacco smoking and targeting smoking cessation efforts to reduce the incidence of depression.

## Data Availability

Data and code required to replicate the analysis findings reported in the article are provided on GitHub at: https://doi.org/10.5281/zenodo.12187311. Primary data from the UK Biobank resource are accessible upon application (https://www.ukbiobank.ac.uk/). The summary level data sets used for the exposures in MR analyses are available online; GSCAN (https://conservancy.umn.edu/items/ca7ed549-636b-41c0-ae79-97c57e266417); Buchwald and colleagues (https://www.ebi.ac.uk/gwas/efotraits/EFO_0007794).

https://www.ukbiobank.ac.uk/

https://conservancy.umn.edu/items/ca7ed549-636b-41c0-ae79-97c57e266417

https://www.ebi.ac.uk/gwas/efotraits/EFO_0007794

## ACKNOWLEDGEMENTS

We would like to thank Rachel Tyndale and her team for their support in the provision of summary statistics. Quality Control filtering of the UK Biobank data was conducted by R. Mitchell, G. Hemani, T. Dudding, L. Corbin, S. Harrison, L. Paternoster as described in the published protocol (doi: 10.5523/bris.1ovaau5sxunp2cv8rcy88688v). The MRC IEU UK Biobank GWAS pipeline was developed by B. Elsworth, R. Mitchell, C. Raistrick, L. Paternoster, G. Hemani, T. Gaunt (doi: 10.5523/bris.pnoat8cxo0u52p6ynfaekeigi). This research has been conducted using the UK Biobank Resource under Application Number 9142. We thank all the contributors to the consortia we have used GWAS results from in our analyses. We would also like to thank the research participants of UK Biobank for making this work possible.

