## Supplementary Materials for "Disentangling the effects of nicotine versus non-nicotine constituents of tobacco smoke on major depressive disorder: A multivariable Mendelian randomization study"

### Supplementary Notes

**Supplementary Note S1.** Deviations from pre-registered analysis plan.

An analysis plan was pre-registered on the Open Science Framework (OSF) on December 18th 2023 (https://osf.io/udqkm). There are two minor deviations from the analysis plan to report.

**(1)** Exclusion of related individuals:

We stated in the pre-registration that we would exclude people from the UK Biobank sample that were related, which we did not do when performing the GWAS of MDD. We pre-specified that our approach to performing the GWAS would be the MRC IEU UK Biobank GWAS pipeline version 2) [1]. This pipeline offers two options to performing GWAS, using either PLINK or BOLT-LMM software, but we did not specify which option we would use. We chose to employ BOLT-LMM, which uses a linear mixed model (LMM) to account for both relatedness and population stratification and allowed for a wider range of individuals to be included in terms of relatedness and ancestry [1,2]. Given that we were running GWAS stratified by smoking status (i.e., reduced sample size) it was more advantageous to use a method which preserved a greater sample size. As such, while not excluding related individuals is technically a deviation from our pre-registered protocol, relatedness is still accounted for in our GWAS of MDD due to the use of BOLT-LMM.

**(2)** Clumping threshold for MVMR analysis:

We stated in the pre-registration that we would perform "additional clumping prior to the MVMR analysis ($r^2$ <0.001, >500kb)". This stated clumping threshold was an error in the pre-registration, as we intended to follow the approach taken in the published paper examining the independent effects of NMR and CPD on physical health outcomes [3], in which the instruments are clumped at ($r^2$ <0.1, >500kb).

**Supplementary Note S2.** Information about the genotyping, quality control, and imputation methods for the UK Biobank data.

This information is taken from the MRC IEU UK Biobank GWAS pipeline (Version 2) recommended paragraphs for publication [1].

The full data release contains the cohort of successfully genotyped samples (n=488,377). 49,979 individuals were genotyped using the UK BiLEVE array and 438,398 using the UK Biobank axiom array. Pre-imputation QC, phasing and imputation are described elsewhere [4]. In brief, prior to phasing, multiallelic SNPs or those with MAF ≤1% were removed. Phasing of genotype data was performed using a modified version of the SHAPEIT2 algorithm [5]. Genotype imputation to a reference set combining the UK10K haplotype and HRC reference panels [6] was performed using IMPUTE2 algorithms [7]. The analyses presented here were restricted to autosomal variants within the HRC site list using a graded filtering with varying imputation quality for different allele frequency ranges. Therefore, rarer genetic variants are required to have a higher imputation INFO score (Info>0.3 for MAF >3%; Info>0.6 for MAF 1-3%; Info>0.8 for MAF 0.5-1%; Info>0.9 for MAF 0.1-0.5%) with MAF and Info scores having been recalculated on an in-house derived 'European' subset [4].

Individuals with sex-mismatch (derived by comparing genetic sex and reported sex) or individuals with sex-chromosome aneuploidy were excluded from the analysis (n=814). We restricted the sample to individuals of 'European' ancestry as defined by an in-house k-means cluster analysis performed using the first 4 principal components provided by UK Biobank in the statistical software environment R. The current analysis includes the largest cluster from this analysis (n=464,708) [4]. To model population structure in the sample we used 143,006 directly genotyped SNPs, obtained after filtering on MAF > 0.01; genotyping rate > 0.015; Hardy-Weinberg equilibrium p-value < 0.0001 and LD pruning to an $r^2$ threshold of 0.1 using PLINKv2.00.

**Supplementary Note S3.** Approach to defining MDD case/control status for GWAS in UK Biobank.

To perform the GWAS of MDD in UK Biobank, we followed the approach outlined by Glanville and colleagues [8] which aims to improve the identification of MDD in UK Biobank using multiple indicators of depression.

We first split the UK Biobank cohort by MHQ participation. For individuals who did not participate in the MHQ, endorsement of five depression phenotypes was calculated ('Help-seeking', 'Self-reported depression', 'Antidepressant usage', 'Depression (Smith)' or 'Hospital (ICD-10) depression'. We then counted the number of measures endorsed by each individual. Detailed information on each phenotype is reported in Supplementary Note **S4**. Cases were defined as individuals endorsing ≥2 depression phenotypes, as the strength of genetic contribution to cases with at least two measures was found to approximate that for CIDI-defined (i.e., gold-standard measure) cases [8].

For individuals who did participate in the MHQ, cases were identified as individuals with a lifetime history of depression from responses to the CIDI depression module. Scoring criteria have been previously defined [9] and are equivalent to the DSM criteria for MDD [9]. Detailed information on the items contributing to the CIDI depression module are reported in Supplementary Note **S4**.

Participants were also screened for five psychosis phenotypes: 'Self-reported psychosis', 'Antipsychotic usage', 'Bipolar (Smith)', 'Hospital (ICD-10) psychosis' and 'Psychosis (MHQ screen)'. Detailed information on these phenotypes are reported in Supplementary Note **S5**. Individuals meeting criteria for any of the psychosis phenotypes were excluded from the analysis (i.e., did not meet case or control criteria).

Controls comprised all UK Biobank participants who did not meet the criteria for depression or psychosis phenotypes, as follows:

**(1)** Did not meet the criteria for any indication of depression i.e., ‘Help-seeking’, ‘Self-reported depression’, ‘Antidepressant usage’, ‘Depression (Smith), ‘Hospital (ICD-10) depression’ or ‘Lifetime depression (MHQ)’; **AND**

**(2)** Did not meet the criteria for any indication of psychosis i.e., ‘Self-reported psychosis’, ‘Antipsychotic usage’, ‘Bipolar (Smith), ‘Hospital (ICD-10) psychosis’ or ‘Psychosis (MHQ)’; **AND**

**(3)** Did not endorse the question: “Have you been diagnosed with one or more of the following mental health problems by a professional, even if you do not have it currently [UK Biobank Field ID 20544] for depression (11 = “Depression”) in the MHQ Section A screening questions.

**Supplementary Note S4.** Derivation of depression phenotypes in UK Biobank.

This section describes the criteria for defining cases for each of the depression phenotypes derived from different sources of phenotypic information in UK Biobank. [ID] refers to the corresponding UK Biobank Field ID, which are browsable via the data showcase platform: https://biobank.ndph.ox.ac.uk/showcase/.

#### Help-seeking

Criteria for defining ‘help-seeking’ cases was endorsing either of the following questions at baseline, or the subsequent repeat assessments:

| Measure | Coding | ID |
|---|---|---|
| “Have you ever seen a general practitioner (GP) for nerves, anxiety, tension or depression?” | 1 = “Yes” | 2090 |
| “Have you ever seen a psychiatrist for nerves, anxiety, tension or depression?” | 1 = “Yes” | 2100 |

#### Self-reported depression

Criteria for defining ‘self-reported depression’ was endorsing “depression” at baseline or the subsequent repeat assessments:

| Measure | Coding | ID |
|---|---|---|
| Code for non-cancer illness. If the participant was uncertain of the type of illness they had had, then they described it to the interviewer (a trained nurse) who attempted to place it within the coding tree. If the illness could not be located in the coding tree then the interviewer entered a free-text description of it. These free-text descriptions were subsequently examined by a doctor and, where possible, matched to entries in the coding tree. Free-text descriptions which could not be matched with very high probability have been marked as "unclassifiable". | 1286 = “depression” | 20002 |

#### Antidepressant usage

Criteria for defining ‘antidepressant usage’ cases was self-reported antidepressant medication at the baseline or the subsequent repeat assessments:

| Measure | Codes | ID |
|---|---|---|

| | | |
|---|---|---|
| Medication Status was obtained via a verbal interview item requesting the names of regular prescription medications that the participants were currently taking. The nurses conducting the interviews did not record medications that were short-term (e.g., 1-week course of antibiotics), historical, or prescribed but not being taken. | 1140879616, 1140921600, 1140879540, 1140867878, 1140916282, 1140909806, 1140867888, 1141152732, 1141180212, 1140879634, 1140867876, 1140882236, 1141190158, 1141200564, 1140867726, 1140879620, 1140867818, 1140879630, 1140879628, 1141151946, 1140867948, 1140867624, 1140867756, 1140867884, 1141151978, 1141152736, 1141201834, 1140867690, 1140867640, 1140867920, 1140867850, 1140879544, 1141200570, 1140867934, 1140867758, 1140867914, 1140867820, 1141151982, 1140882244, 1140879556, 1140867852, 1140867860, 1140917460, 1140867938, 1140867856, 1140867922, 1140910820, 1140882312, 1140867944, 1140867784, 1140867812, 1140867668 | 20003 |

**Depression (Smith)**

Criteria for defining 'Depression (Smith)' cases was meeting the criteria for one of three depression phenotypes at baseline, defined previously by Smith and colleagues [10] and described in detail under resource 158722:

| **Measure** | **Codes** | **ID** |
|---|---|---|
| Single Probable major depression episode [10] | 5 = Single Probable major depression episode | 20126 |
| Probable recurrent major depression (moderate) [10] | 4 = Probable recurrent major depression (moderate) | 20126 |
| Probable Recurrent major depression (severe) [10] | 3 = Probable Recurrent major depression (severe) | 20126 |

**Hospital (ICD-10) depression**

Criteria for ‘Hospital (ICD-10)’ cases was being admitted for hospital inpatient care with a diagnosis (either primary or secondary) for major depression ([ICD-10] = F32.X and F33.X) between April 1997 and November 2023:

| **Measure** | **Codes** | **ID** |
|---|---|---|
| Depressive episode | F32, F320, F321, F322, F323, F328, F329 | 41202<br>41204 |
| Recurrent depressive disorder | F33, F330, F331, F332, F334, F338, F339 | 41202<br>41204 |

**Lifetime depression (MHQ)**

Criteria for defining ‘Lifetime Depression (MHQ) cases is detailed below. These mirror criteria defined by Davis and colleagues [9] for identifying individuals with a lifetime history of depression, and mirrors CIDI-SF criteria for lifetime depression. Participants must have endorsed: (i) at least one of the two ‘core symptoms’; **AND** (ii) a score above threshold on the ‘threshold items’; **AND** (iii) experiencing ≥5 symptoms (including core) during ‘worst episode of depression’:

| **Measure** | **Codes** | **ID** |
|---|---|---|
| Core symptoms:<br>• “Have you ever had a time in your life when you felt sad, blue, or depressed for two weeks or more in a row?”;<br>• “Have you ever had a time in your life lasting two weeks or more when you lost interest in most things like hobbies, work, or activities that usually give you pleasure?” | 1 = “Yes” | 20446<br>20441 |
| Threshold items:<br>“Please think of the two-week period in your life when your feelings of depression or loss of interest were the worst.”<br>• “How much of the day did these feelings usually last?” (>2);<br>• “(How often) did you feel this way?” (>1)<br>• “Think about your roles at the time of this episode, including study/employment, childcare and housework, leisure pursuits. How much did these problems interfere with your life or activites?” (>1) | 2 = “About half of the day”<br>1 = “Less often”<br>1 = “A little” | 20436<br>20439<br>20440 |
| Symptoms during worst episode of depression: | | 20449 |

| | | |
|---|---|---|
| "Please think of the two-week period in your life when your feelings of depression or loss of interest were the worst"<br>• "Did you feel more tired out or low on energy than is usual for you?"<br>• "Did you gain or lose weight without trying or did you stay about the same weight?"<br>• "Did your sleep change?"<br>• "Did you have a lot more trouble concentrating than usual?"<br>• "People sometimes feel down on themselves, no good, worthless. Did you feel this way?"<br>• "Did you think a lot about death – either your own, someone else's or death in general?" | <br>1 = "Yes"<br>1 = "Gained weight"; 2 = "Lost weight"; 3 = "Both gained and lost some weight"<br>1 = "Yes"<br>1 = "Yes"<br>1 = "Yes"<br>1 = "Yes" | 20536<br>20532<br>20435<br>20450<br>20437 |

**Supplementary Note S5.** Derivation of psychosis phenotypes in UK Biobank.

This describes the criteria for defining cases for each of the psychosis phenotypes derived from different sources of phenotypic information in UK Biobank. [ID] refers to the corresponding UK Biobank Field ID, which are browsable via the data showcase platform: https://biobank.ndph.ox.ac.uk/showcase/.

#### Self-reported psychosis

Criteria for defining 'self-reported psychosis was endorsing "schizophrenia" or "mania/bipolar disorder/manic depression" at baseline or the subsequent repeat assessments:

| Measure | Coding | ID |
|---|---|---|
| Code for non-cancer illness. If the participant was uncertain of the type of illness they had had, then they described it to the interviewer (a trained nurse) who attempted to place it within the coding tree. If the illness could not be located in the coding tree then the interviewer entered a free-text description of it. These free-text descriptions were subsequently examined by a doctor and, where possible, matched to entries in the coding tree. Free-text descriptions which could not be matched with very high probability have been marked as "unclassifiable". | 1289 = "schizophrenia"<br>1291 = "mania/bipolar disorder/manic depression" | 20002 |

#### Antipsychotic usage

Criteria for defining 'antidepressant usage' cases was self-reported antipsychotic medication at the baseline or the subsequent repeat assessments:

| Measure | Codes | ID |
|---|---|---|
| Medication Status was obtained via a verbal interview item requesting the names of regular prescription medications that the participants were currently taking. The nurses conducting the interviews did not record medications that were short-term (e.g., 1-week course of antibiotics), historical, or prescribed but not being taken. | 1140868170, 1140928916, 1141152848, 1140867444, 1140879658, 1140868120, 1141153490, 1140867304, 1141152860, 1140867168, 1141195974, 1140867244, 1140867152, 1140909800, 1140867420, 1140879746, 1141177762, 1140867456, | 20003 |

| | |
|---|---|
| | 1140867952, 1140867150, 1141167976, 1140882100, 1140867342, 1140863416, 1141202024, 1140882098, 1140867184, 1140867092, 1140882320, 1140910358, 1140867208, 1140909802, 1140867134, 1140867306, 1140867210, 1140867398, 1140867078, 1140867218, 1141201792, 1141200458, 1140867136, 1140879750, 1140867180, 1140867546, 1140928260, 1140927956 |

**Bipolar (Smith)**

Criteria for defining 'Bipolar (Smith)' cases was meeting the criteria for one of two bipolar phenotypes at baseline, defined previously by Smith and colleagues [10] and described in detail under resource 158722:

| **Measure** | **Codes** | **ID** |
|---|---|---|
| Bipolar Type I (Mania) [10] | 1 = Bipolar Type I (Mania) | 20126 |
| Bipolar Type II (Hypomania) [10] | 2= Bipolar Type II (Hypomania) | 20126 |

**Hospital (ICD-10) psychosis**

Criteria for 'Hospital (ICD-10)' cases was being admitted for hospital inpatient care with a diagnosis (either primary or secondary) for a psychotic disorder between April 1997 and November 2023:

| **Measure** | **Codes** | **ID** |
|---|---|---|
| Schizophrenia, schizotypal and delusional disorders | F20, F200, F201, F202, F203, F204, F205, F206, F208, F209, F21, F22, F220, F228, F229, F23, F230, F231, F232, F233, F238, F239, F24, F25, F250, F251, F252, F258, F259, F28, F29 | 41202 41204 |
| Mood [affective] disorders (excluding Depression codes F32-F33) | F30, F300, F301, F302, F308, F309, F31, F310, F311, F312, F313, F314, F315, F316, F317, | 41202 41204 |

| | |
|---|---|
| | F318, F319, F34, F340, F341, F348, F349, F38, F380, F381, F388, F39 |

**Psychosis (MHQ)**

Criteria for 'Psychosis (MHQ)' cases was endorsing "schizophrenia", "any other type of psychosis or psychotic illness" or "mania/hypomania/bipolar/manic-depression" in response to a screening question in MHQ Section A:

| **Measure** | **Codes** | **ID** |
|---|---|---|
| "Have you been diagnosed with one or more of the following mental health problems by a professional, even if you don't have it currently?" | 2 = "Schizophrenia"<br>3 = "Any other type of psychosis or psychotic illness"<br>10 = "Mania, hypomania, bipolar or manic-depression" | 20544 |

**Supplementary Note S6.** Additional details regarding population stratification assessment, harmonisation, clumping, univariable MR methods and assumptions of MR.

**SNP-based heritability:**

SNP-based heritabilities (Supplementary Table **S1**) were calculated using linkage disequilibrium score regression (LDSC v1.0.1) [11,12] using the UK Biobank GWAS summary statistics and pre-computed linkage disequilibrium scores for each SNP calculated based on individuals of European ancestry from using 1000 Genomes European data [13]. LD scores were filtered to HapMap3 SNPs as these are well-imputed in most studies [14]. SNP-based heritabilities were transformed to the liability scale using lifetime risk of 15% [8,15].

Summary statistics were imported into R version 4.3.1 to create QQ plots (Supplementary Figure **S5**) using the package *fastman* [16]. Overall, low LD score intercepts and ratios (Supplementary Table **S1**) suggested that genomic inflation observed in the QQ plots was not driven by population stratification.

**Harmonisation and clumping:**

We used the *TwoSampleMR* R package [17] *harmonise_data* function to harmonise the exposure and outcome datasets. Palindromic SNPs were only excluded if their allele frequency could not be used to infer which strand was positive (i.e., action =2). Two SNPs associated with smoking heaviness (i.e., CPD) were removed due to having intermediate allele frequencies (rs1737894, rs28438420).

Two SNPs were associated with both NMR and CPD (rs56113850, rs117824460), and one SNP associated with CPD was not available in the NMR GWAS dataset (rs4886550). In cases where SNPs were missing in the either exposure dataset, we used the *LDproxy* function from the LDlinkR package [18] to identify proxy SNPs with a minimum linkage disequilibrium ($R^2$) of 0.8. All SNPs available in the exposure GWAS datasets were available in the MDD GWAS dataset. The p-value for the SNP-exposure effect size for one SNP in the NMR instrument (rs117090198) was unusually high prior to conditional analysis in the published GWAS ([19]; Supplementary Information). As the authors could not provide a definitive explanation for this, this SNP was removed to reduced heterogeneity.

In MVMR analyses, all SNPs included in the model should be independent of each other (i.e., SNPs associated with NMR must also be independent of the SNPs associated with CPD, and vice versa) [20]. To ensure overall independence, the full list of SNPs (N = 59) were clumped (LD $R^2 < 0.1$, >500kb). Considering the limited number of SNPs associated with the NMR, compare to the CPD instrument, SNPs associated with CPD were dropped from the analysis to preserve instrument strength (i.e., no NMR SNPs were dropped during the clumping stage).

**Brief description of MR and key assumptions:**

MR is an analytical method which uses genetic variants (typically Single Nucleotide Polymorphisms [SNPs]) as instrumental variables for exposures of interest. The MR approach draws on Mendel's first and second laws of genetic inheritance: the law of segregation and the law of independent assortment [21]. The law of segregation suggests that each of the parent's two copies of a given section of DNA (i.e., allele) has an equal chance of being inherited, therefore offspring randomly inherit one allele from their mother and one allele from their father [22,23]. The law of independent assortment suggests that the two alleles are inherited independently of each other, except in regions of the genome that are genetically linked in the DNA of the parents [22,23]. Furthermore, genotype is determined at conception and germline DNA is not modified subsequently [22]. As such, results from MR studies should be more robust to confounding and reverse causation bias than traditional observational studies.

However, for genetic variants to be used as instrumental variables (IVs) they must satisfy several conditions or assumptions: (1) the IV is associated with the exposure (i.e., relevance assumption), (2) there are no causes of the IV that also influence the outcome through mechanisms other than the exposure of interest (i.e., independence assumption), (3) the IV does not affect the outcome other than through the exposure, and does not affect any other trait that has a downstream effect of the outcome of interest (i.e., exclusion restriction assumption) [22,23]. The IV conditions can be expressed with directed acyclic graphs, or DAGs; see below (solid red lines depict effects which must exist, dashed lines depict effects which must not exist, **G** is the IV (i.e., genetic variant(s) to proxy exposure), **U** represents unobserved confounders; adapted from [23]).

**IV1:**

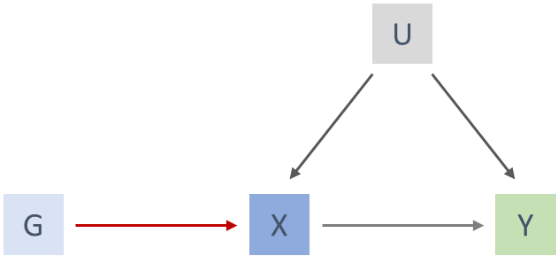

**IV2:**

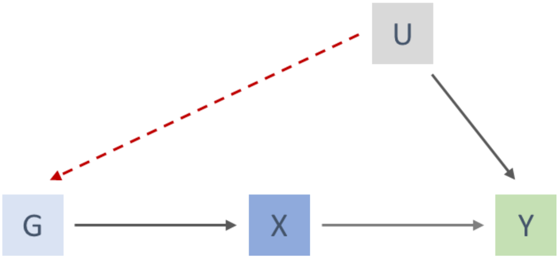

**IV3:**

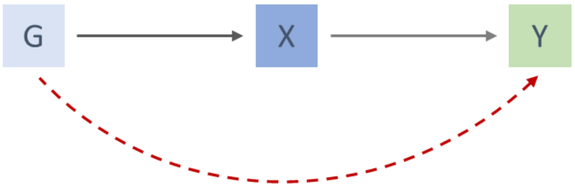

The first IV condition can be tested empirically, through generating an F statistic which tests the associations between the SNPs and the exposure. As a general rule-of-thumb, F>10 has been used to indicate the level of bias due to weak instruments is likely to be small. The second and third IV conditions cannot be proved to be true but can be disproved (i.e., absence of evidence for invalid IVs) [23]. As genetic variants are fixed at conception, conventional confounders (e.g., age, sex) cannot influence them; however, confounding of the IVs with the outcome can be induced by population stratification, dynastic effects and assortative mating [23], which violates IV2. The use of negative controls can be used to explore violations of IV2 due to population stratification [24]. Violations of IV3 can be caused by pleiotropy (i.e., genetic variants have effects on multiple phenotypes). There are many MR methods available which are robust to different forms of pleiotropy, which can be used to explore how sensitive results are to the assumption of no pleiotropy [25].

**Univariable MR methods:**

This study used four complementary univariable MR methods: inverse variance weighted (IVW), MR-Egger, weighted median-based estimation and weighted modal-based estimation [26–29]. This follows recommendations for performing two-sample, or 'summary-level' MR with a variety of robust methods which rely on different assumptions regarding pleiotropy [25]. The IVW method was used as our primary estimator; the IVW statistic is a weighted mean of the ratio estimates for individual SNP instruments (i.e., Wald ratios, calculated by dividing the SNP-outcome association by the SNP exposure association) and forces the intercept through zero thus assuming no horizontal pleiotropy [26]. As such, if only one genetic variant is not a valid IV the estimator will be biased [25]. The MR-Egger method instead assumes a non-zero intercept to account for pleiotropy and provides a consistent causal estimate when the Instrument Strength Independent of Direct Effect (InSIDE) assumption is met which states that SNPs' pleiotropic effects on the outcome are independent of the SNPs association with the exposure [27]. As the intercept is not constrained to pass through zero, it provides an estimate of the directional pleiotropic effect [25]. Reliability of the Egger regression estimate can be assessed using the $I^2_{GX}$ statistic, which indicates expected regression dilution bias and tests violation of the 'NO Measurement Error' (NOME) assumption, with values >0.9 suggesting dilution does not materially affect the MR-Egger analyses [30]. The weighted-median method instead of taking the weighted mean of the ratio estimates (i.e., IVW method) takes the median from a distribution of the ratio estimates in which SNPs with more precise ratio estimates receive more weight [28] and provides an unbiased estimate if up to 50% of the weight comes from variants that are valid IVs or 'majority valid' [25]. The weighted-mode method will provide a consistent estimate under the 'plurality valid' assumption, such that most common causal effect estimates come from valid SNPs even if the majority of SNPs are not valid [25,29].

**Supplementary Note S7.** Results of the univariable and multivariable MR analyses exploring the effect of NMR and CPD on MDD among current and former smokers.

We have focused on the results among ever smokers rather than current or former smokers as the outcome is binary lifetime MDD, and due to the average age of onset of MDD (e.g., 31 years) [31] we cannot guarantee that MDD develops subsequent to changes in self-reported smoking status reported in the middle-age to older-adult sample (i.e., 38-70 years) of UK Biobank. Whereas the average age of starting smoking (i.e., ever smoking) is prior to the average age of onset of MDD, with ~90% of individuals initiating smoking before 20-24 years of age [32]. As such the results are more interpretable among ever smokers.

Among current smokers, the MR-IVW results indicate no clear evidence for a causal effect of NMR on MDD risk ($OR^{IVW}$ = 1.00, 95% CI 0.97 – 1.03; p = 0.975), supported by the MR-Egger, weighted median and weighted mode results ($OR^{EGG}$ = 1.01, 95% CI 0.95 – 1.08, *p* = 0.701; $OR^{MED}$ = 1.01, 95% CI 0.97 – 1.04, *p* = 0.712; $OR^{MOD}$ = 1.01, 95% CI 0.97 – 1.05, *p* = 0.687; respectively). The MR-IVW results provide weak evidence for a causal effect of CPD on MDD risk ($OR^{IVW}$ = 1.14, 95% CI 1.00 – 1.30; p = 0.056), supported by the MR-Egger, weighted median and weighted mode results ($OR^{EGG}$ = 1.19, 95% CI 0.97 – 1.46, *p* = 0.095; $OR^{MED}$ = 1.15, 95% CI 0.96 – 1.38, *p* = 0.140; $OR^{MOD}$ = 1.18, 95% CI 1.00 – 1.40, *p* = 0.056; respectively). The MVMR-IVW results provide no clear evidence for an independent causal effect of NMR on MDD risk ($OR^{IVW}$ = 0.99, 95% CI 0.96 – 1.03, p = 0.721) supported by the MVMR-Egger analyses ($OR^{EGG}$ = 1.00, 95% CI 0.96 – 1.03, p = 0.782). The MVMR-IVW results indicated no clear evidence that increased smoking heaviness increases MDD risk ($OR^{IVW}$ = 1.10, 95% CI 0.87 – 1.40, p = 0.417), supported by the MVMR-Egger analyses ($OR^{EGG}$ = 1.20, 95% CI 0.81 – 1.77, p = 0.367). There was no clear evidence of heterogeneity indicated by the Cochran Q statistic, where values were less than the number of SNPs for all analyses, and Egger intercepts were close to the null (Supplementary Table **S3**) suggesting low likelihood of bias due to directional pleiotropy.

Among current smokers, Steiger filtering identified 3/53 (i.e., 5.66%) of CPD SNPs which explained more variance in the outcome. Univariable MR was repeated with these SNPs excluded. Effect sizes and direction of effect were very similar pre- and post-filtering across primary ($OR^{IVW}$ = 1.13, 95% CI 0.99 – 1.30; p = 0.067) and sensitivity analyses ($OR^{EGG}$ = 1.20, 95% CI 0.98 – 1.48, *p* = 0.088; $OR^{MED}$ = 1.15, 95% CI 0.96 – 1.37, *p* = 0.133; $OR^{MOD}$ = 1.17, 95% CI 1.01 – 1.37, *p* = 0.048) suggesting low risk of bias due to reverse causation.

Among former smokers, the MR-IVW results indicate no clear evidence for a causal effect of NMR on MDD risk ($OR^{IVW}$ = 0.99, 95% CI 0.98 – 1.01; p = 0.206), supported by the MR-Egger, weighted median and weighted mode results ($OR^{EGG}$ = 1.00, 95% CI 0.96 – 1.03, *p* = 0.899; $OR^{MED}$ = 0.99, 95% CI 0.97 – 1.01, *p* = 0.298; $OR^{MOD}$ = 0.99, 95% CI 0.97 – 1.01, *p* = 0.391; respectively). The MR-IVW results provide evidence for a causal effect of CPD on MDD risk ($OR^{IVW}$ = 1.13, 95% CI 1.02 – 1.24; p = 0.048), supported by the MR-Egger, weighted median and weighted mode results ($OR^{EGG}$ = 1.07, 95% CI 0.93 – 1.23, *p* = 0.362; $OR^{MED}$ = 1.11, 95% CI

0.99 – 1.23, *p* = 0.062; $OR^{MOD}$ = 1.12, 95% CI 1.02 – 1.23, *p* = 0.024; respectively). The MVMR-IVW results provide no clear evidence for an independent causal effect of NMR on MDD risk ($OR^{IVW}$ = 0.99, 95% CI 0.98 – 1.01, p = 0.421) supported by the MVMR-Egger analyses ($OR^{EGG}$ = 1.00, 95% CI 0.96 – 1.03, p = 0.899). The MVMR-IVW results indicated evidence that increased smoking heaviness increases MDD risk ($OR^{IVW}$ = 1.20, 95% CI 1.02 – 1.41, p = 0.034), supported by the MVMR-Egger analyses ($OR^{EGG}$ = 1.11, 95% CI 0.84 – 1.45, p = 0.470). There was evidence of heterogeneity indicated by the Cochran Q statistic for the univariable smoking heaviness analyses, and the multivariable MR analyses (Supplementary Table **S3**). However, all Egger intercepts were close to the null (Supplementary Table **S3**), suggesting low likelihood of bias due to directional pleiotropy.

**Supplementary Figure S1.** Flowchart of GWAS of MDD in UK Biobank among ever smokers.

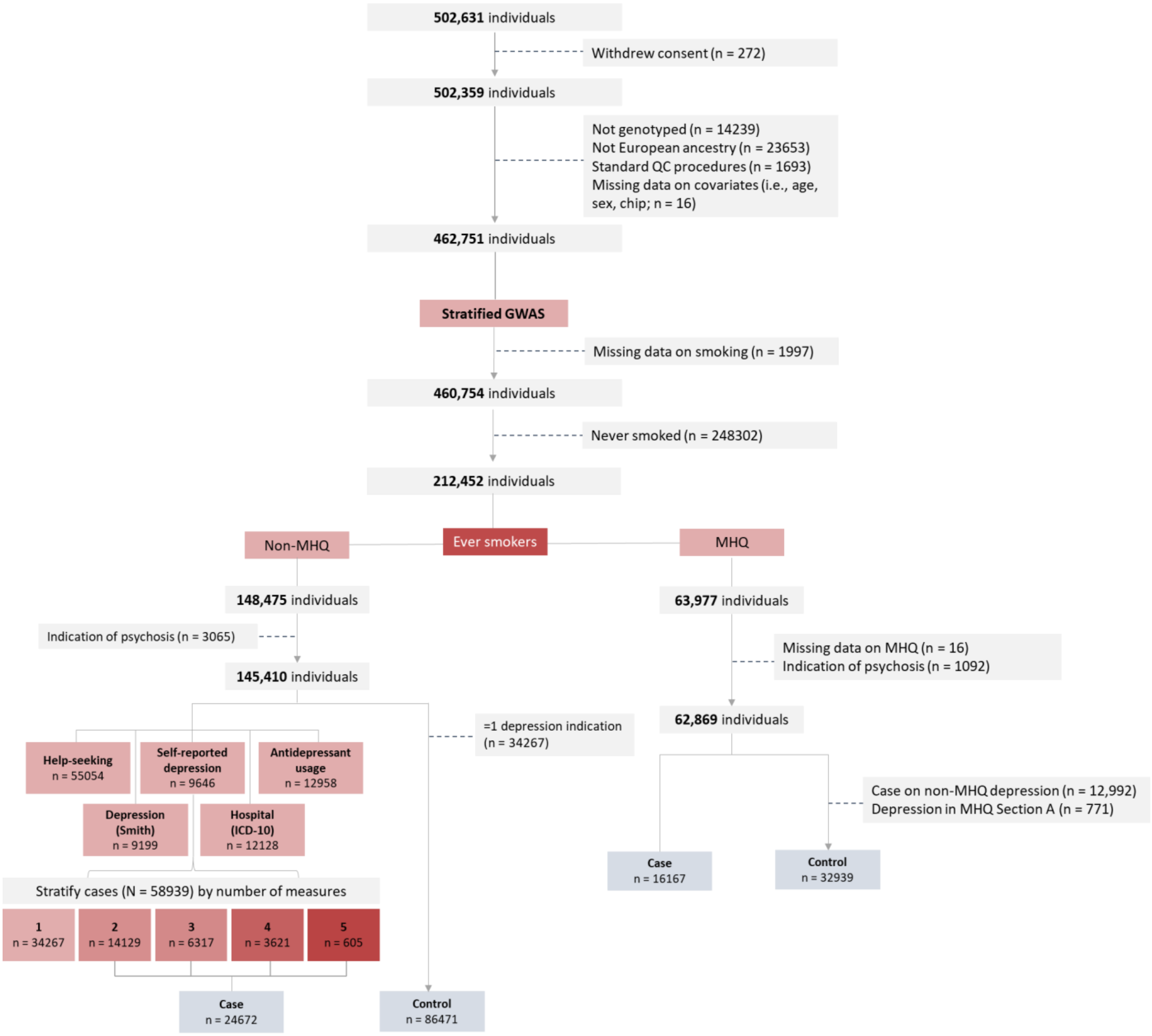

**Supplementary Figure S2.** Flowchart of GWAS of MDD in UK Biobank among never smokers.

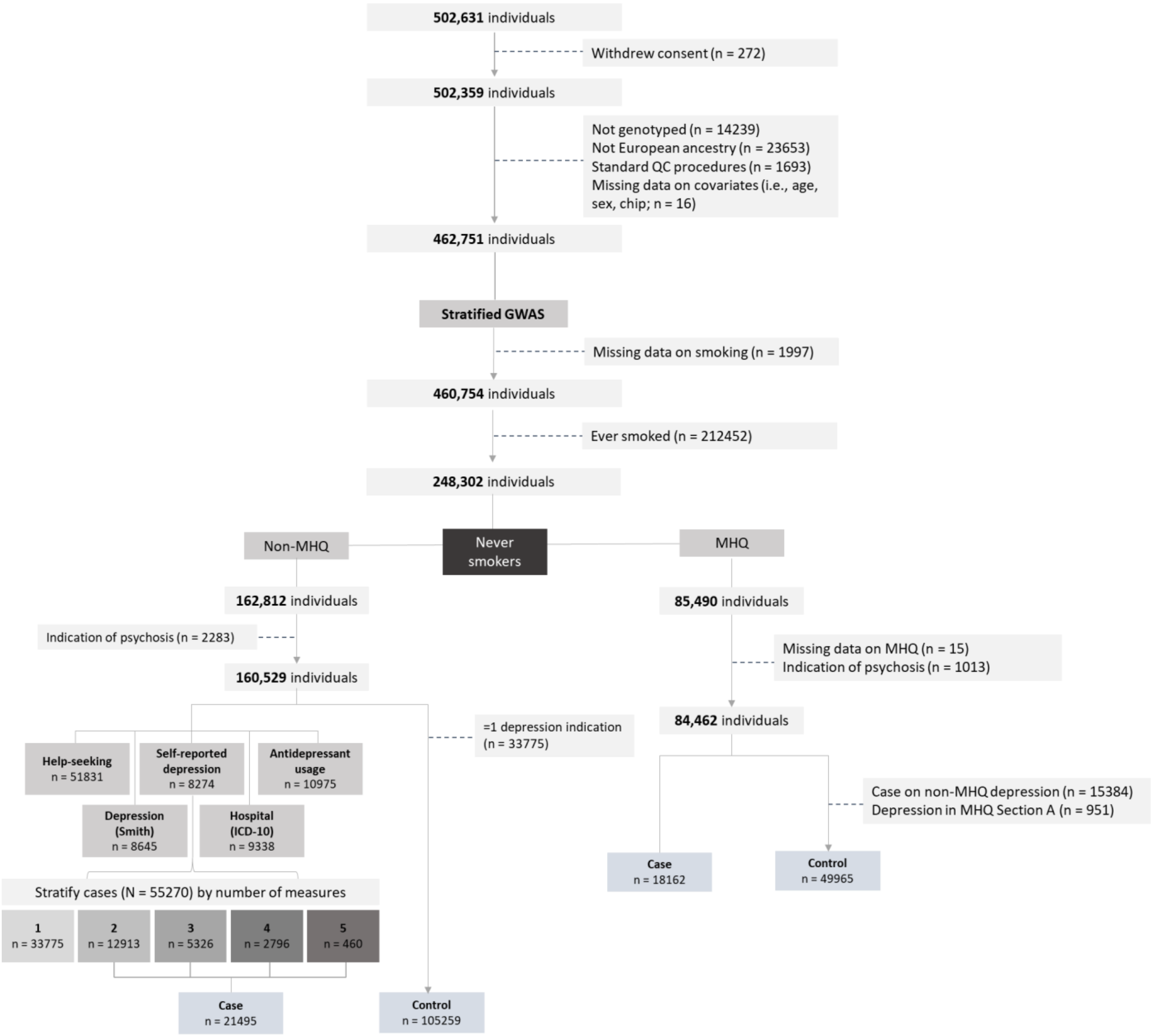

**Supplementary Figure S3.** Flowchart of GWAS of MDD in UK Biobank among current smokers.

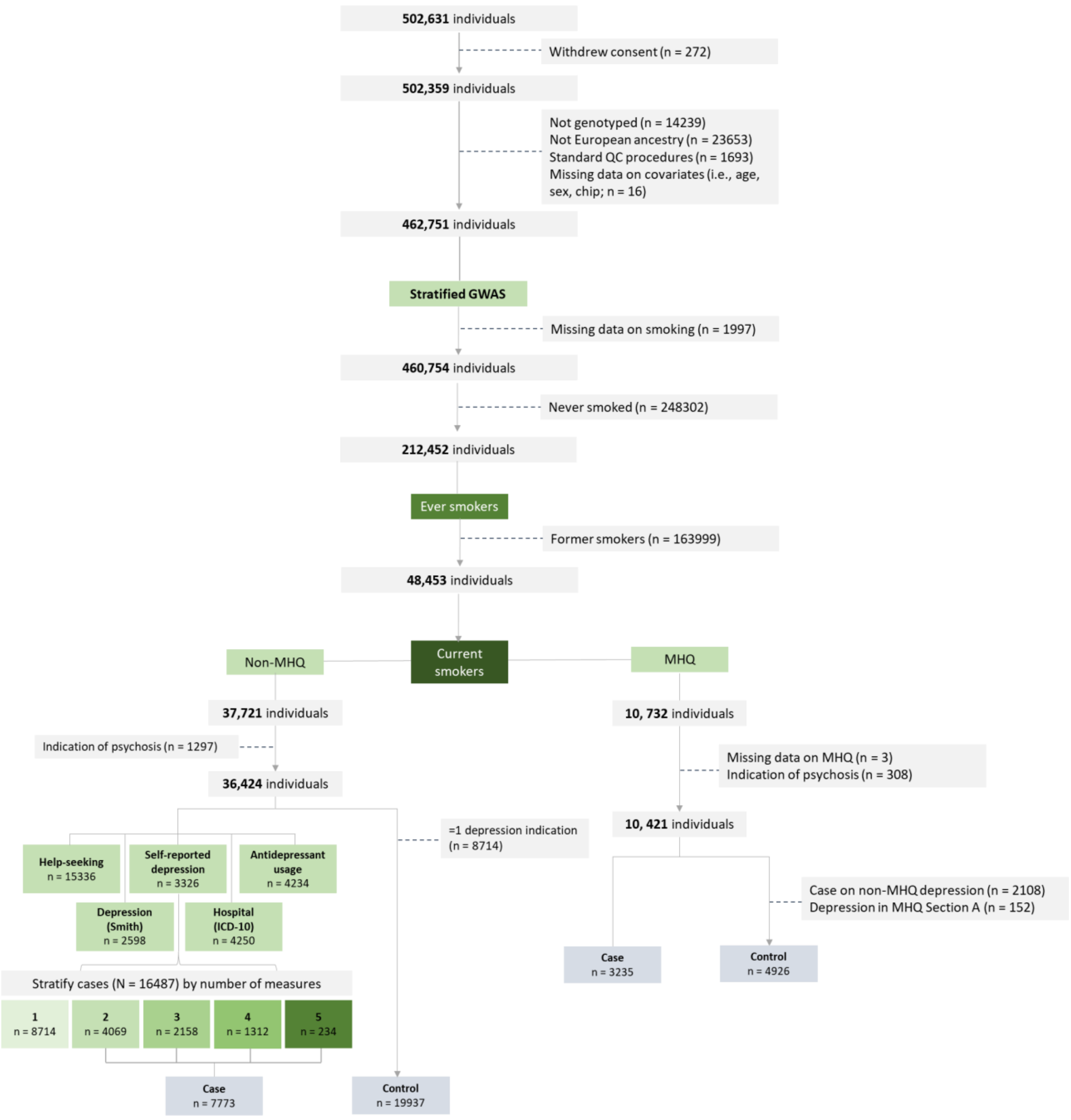

**Supplementary Figure S4.** Flowchart of GWAS of MDD in UK Biobank among former smokers.

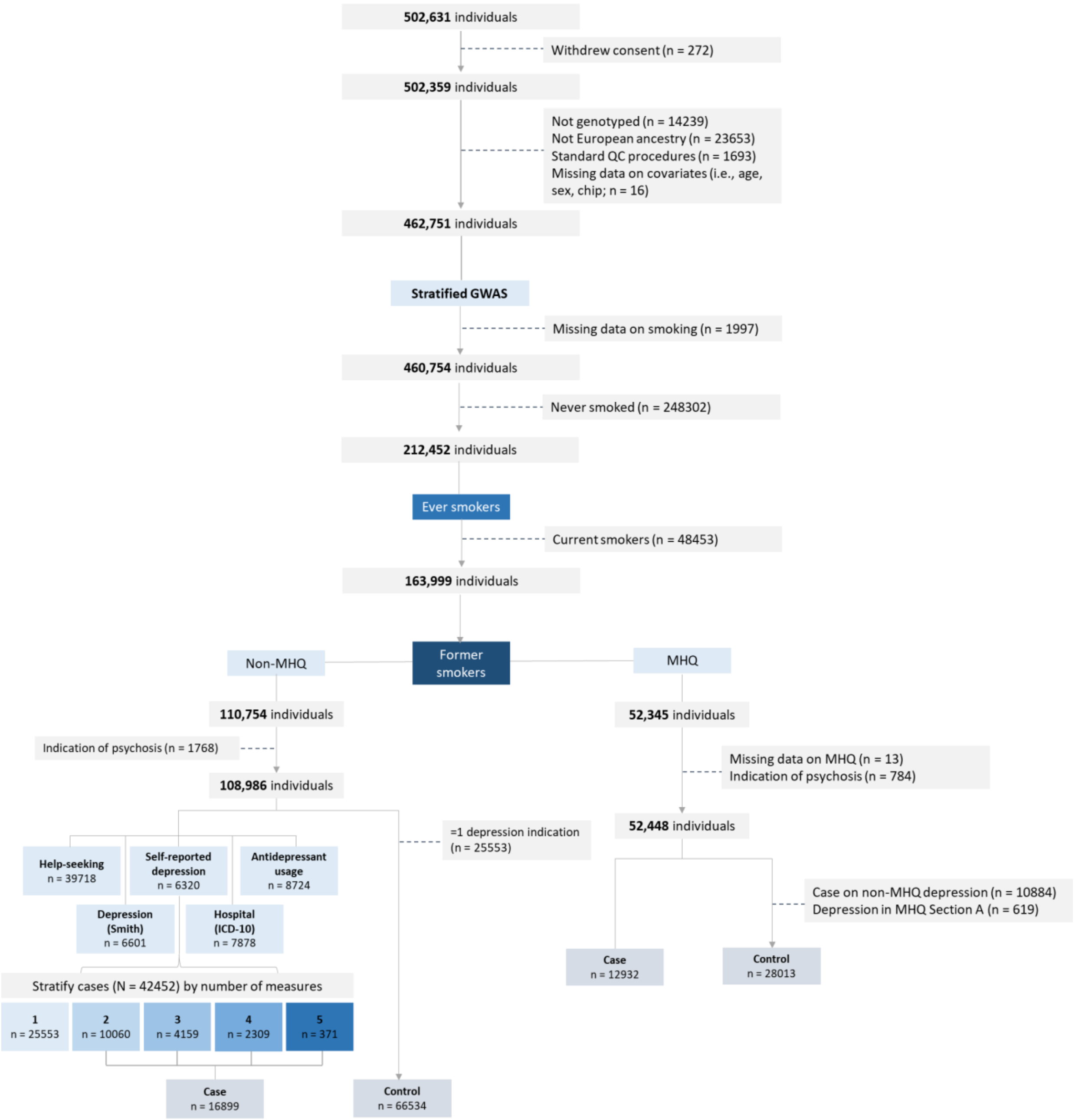

**Supplementary Figure S5**. QQ plots for GWAS of MDD in UK Biobank among (A) ever smokers, (B) never smokers, (C) current smokers and (D) former smokers.

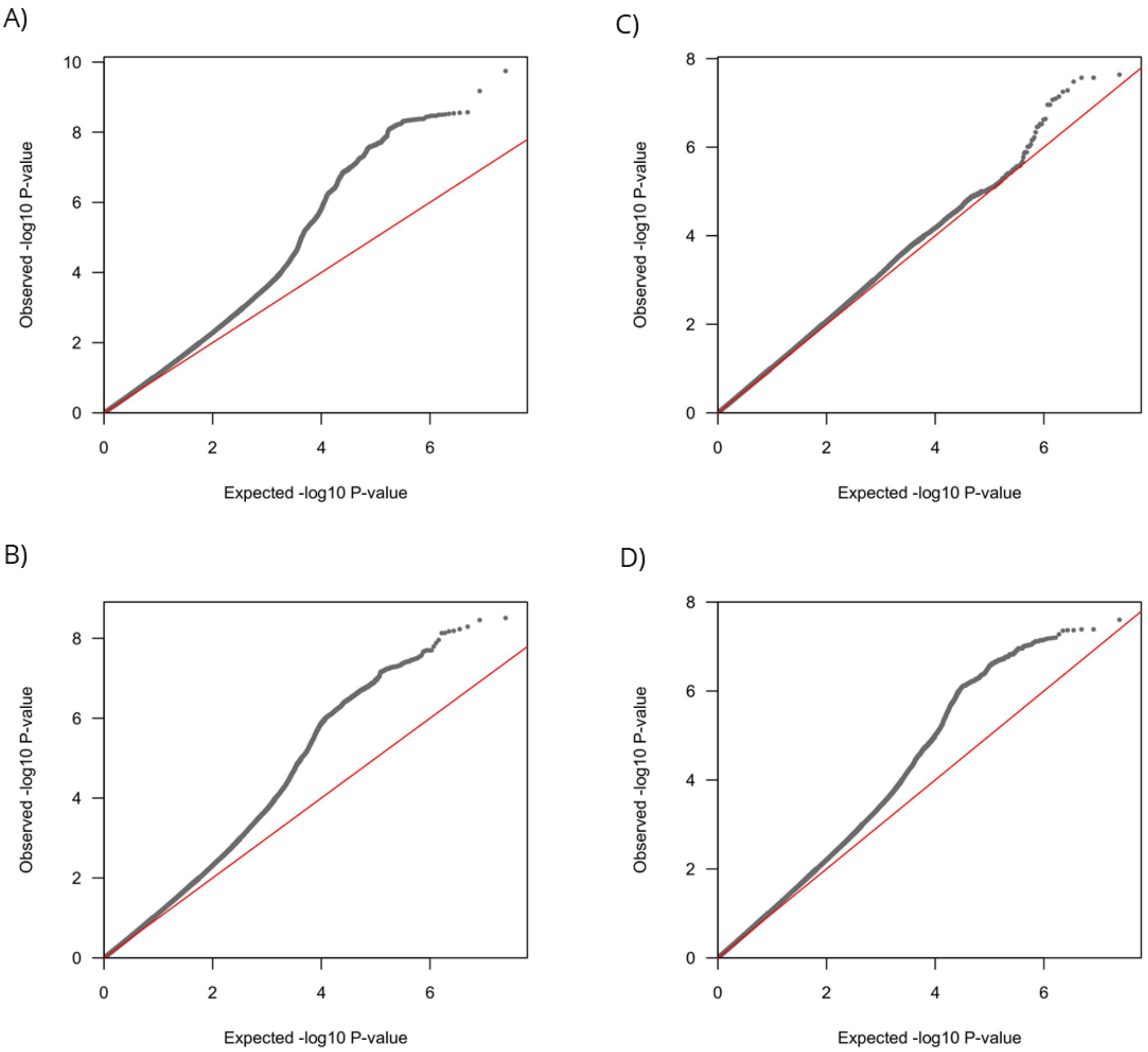

**Supplementary Figure S6.** Scatter plot of associations of the selected variants for NMR with MDD among ever smokers.

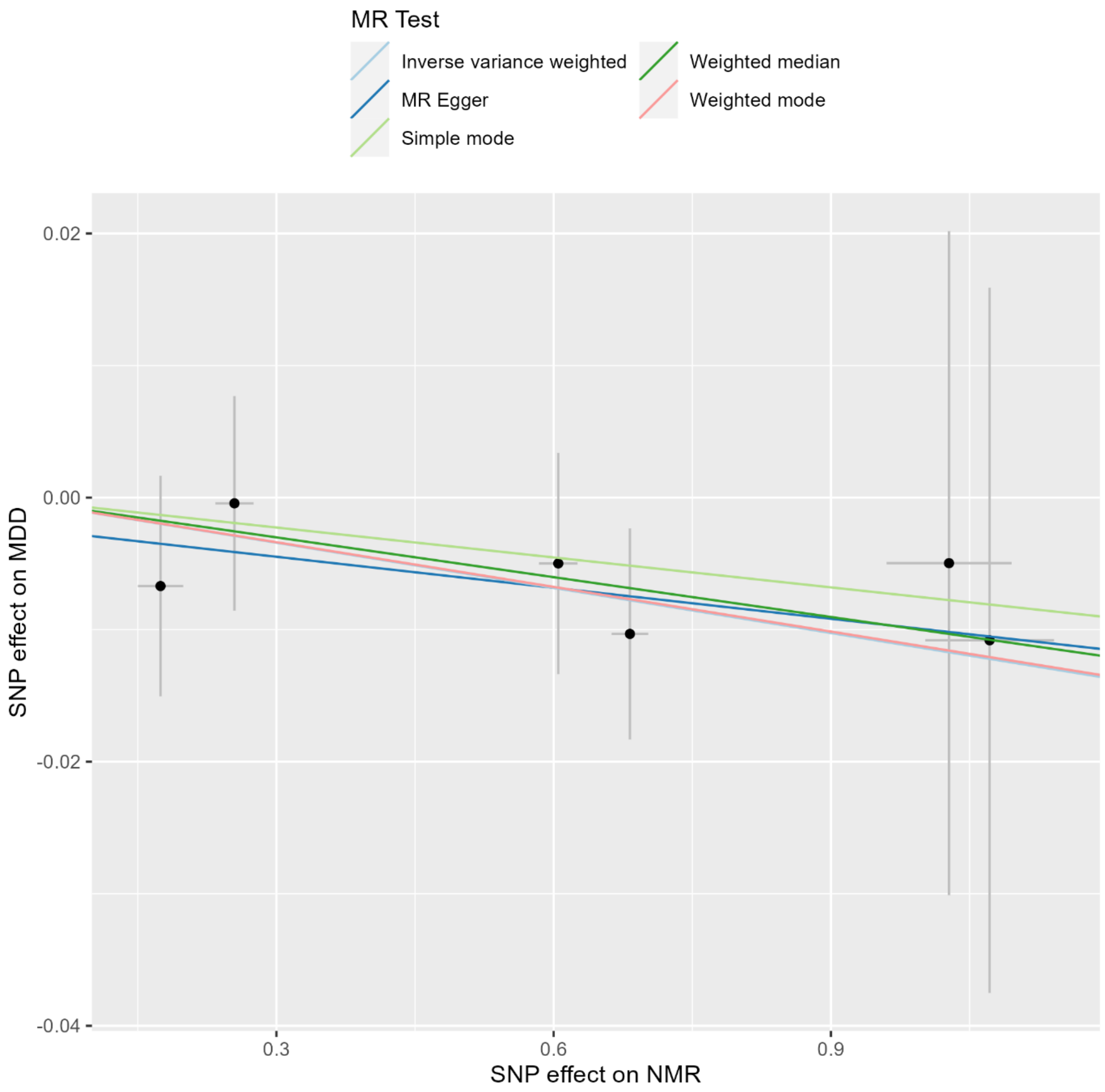

**Supplementary Figure S7.** Scatter plot of associations of the selected variants for CPD with MDD among ever smokers.

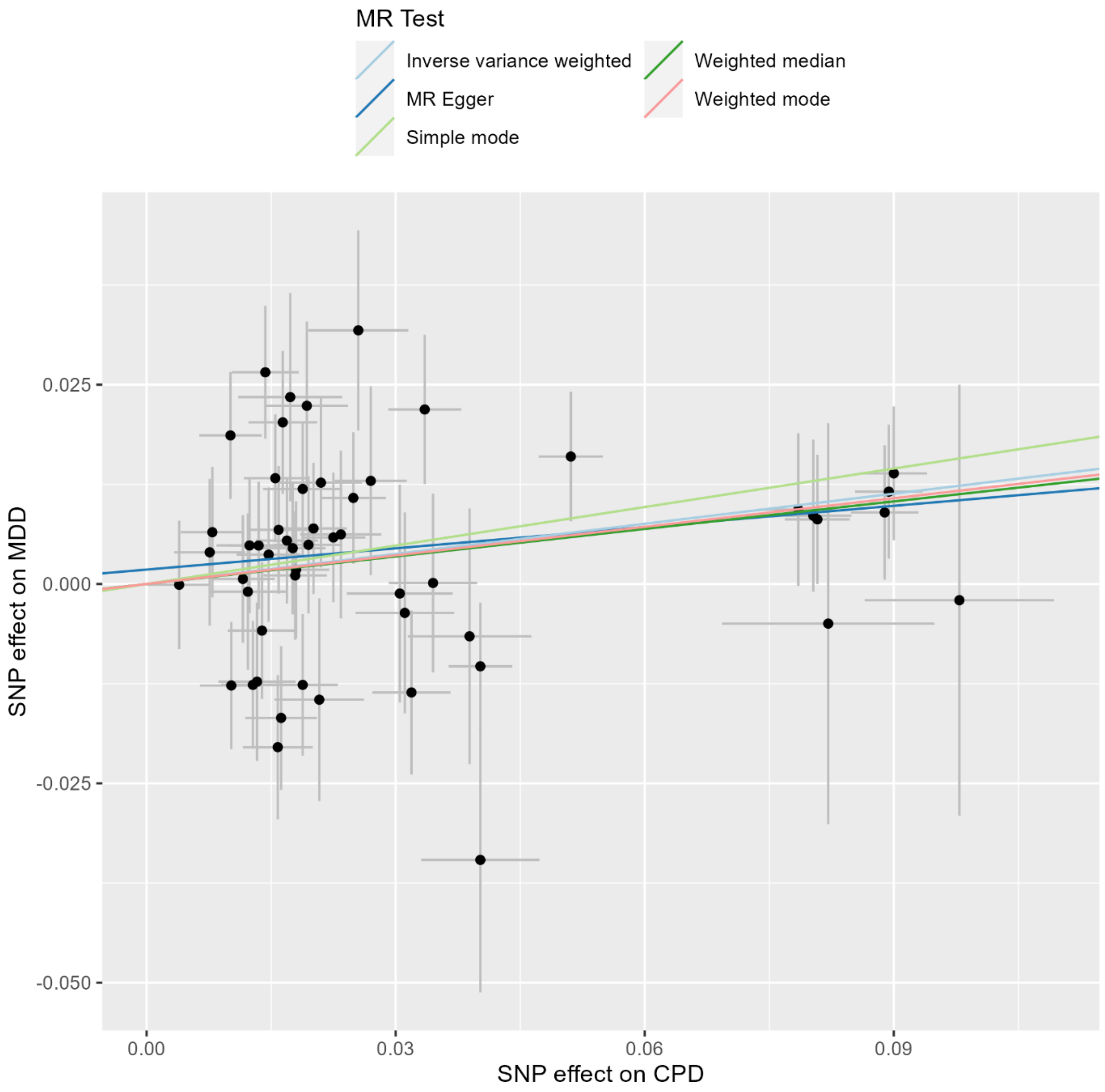

**Supplementary Figure S8.** Forest plot displaying the effect of NMR and smoking heaviness on MDD on MDD: univariable (MR) and multivariable MR (MVMR) results among never smokers.

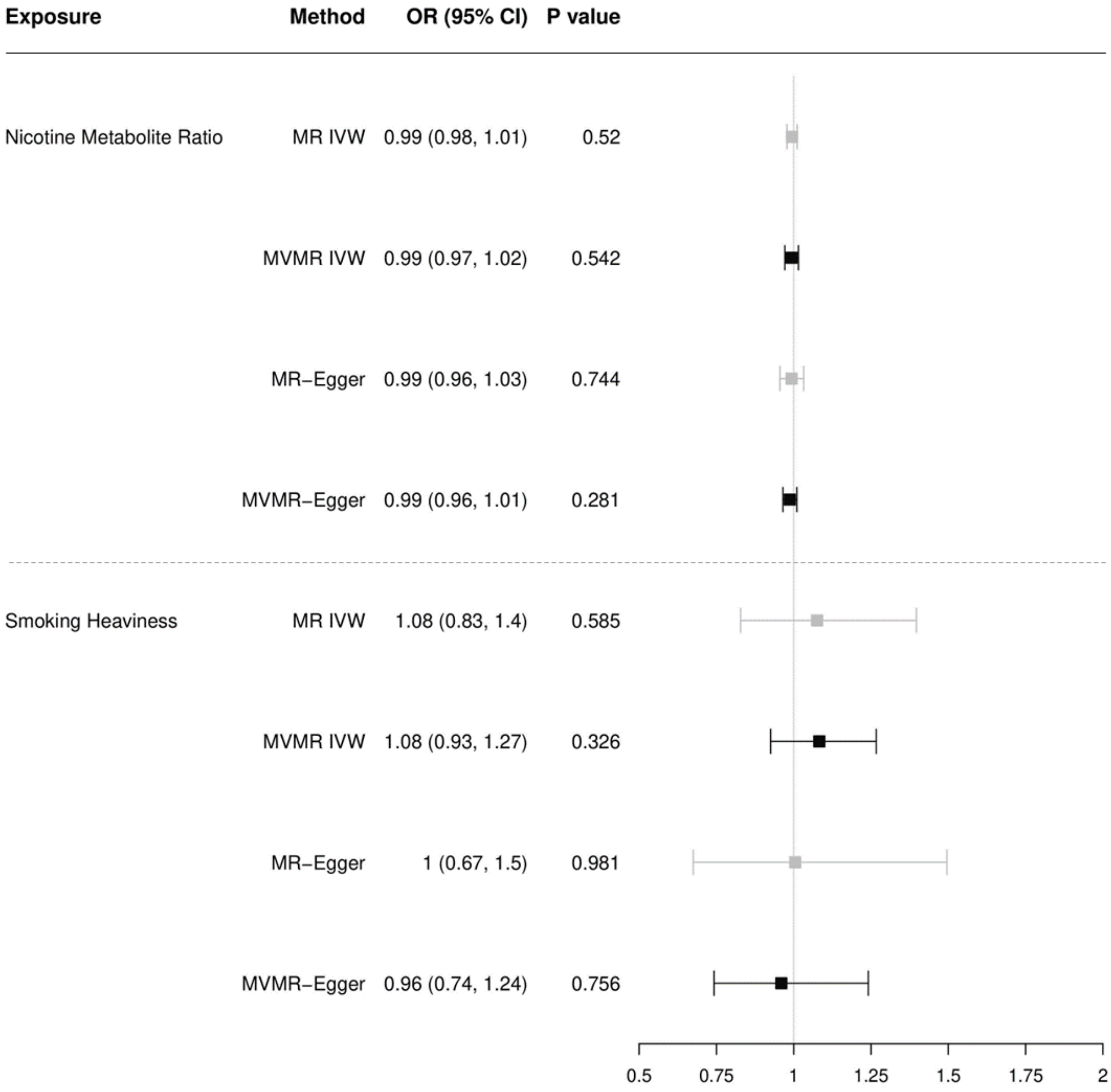

**Supplementary Figure S9.** Forest plot displaying the effect of NMR and smoking heaviness on MDD on MDD: univariable (MR) and multivariable MR (MVMR) results among current smokers.

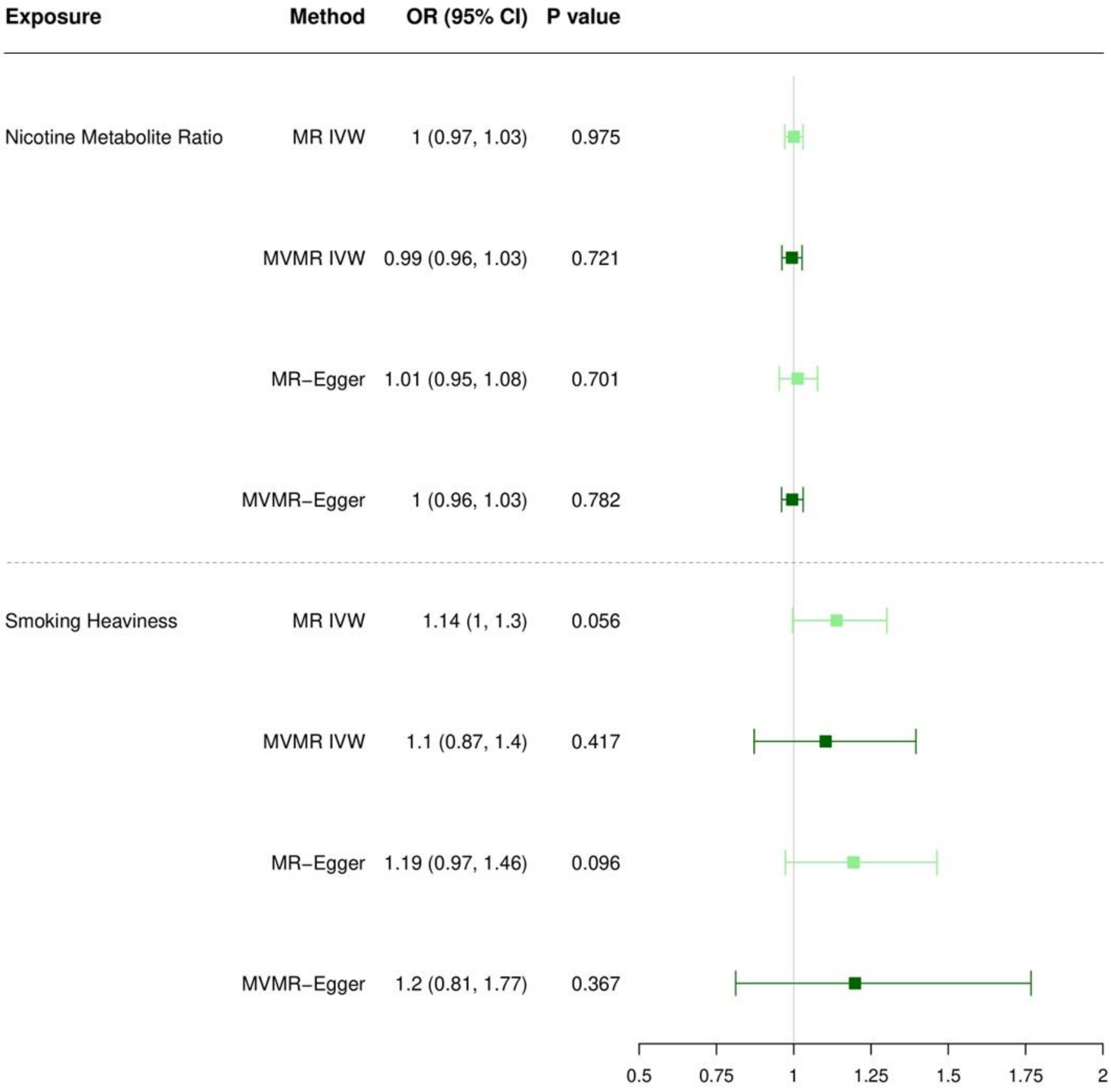

**Supplementary Figure S10.** Forest plot displaying the effect of NMR and smoking heaviness on MDD on MDD: univariable (MR) and multivariable MR (MVMR) results among former smokers.

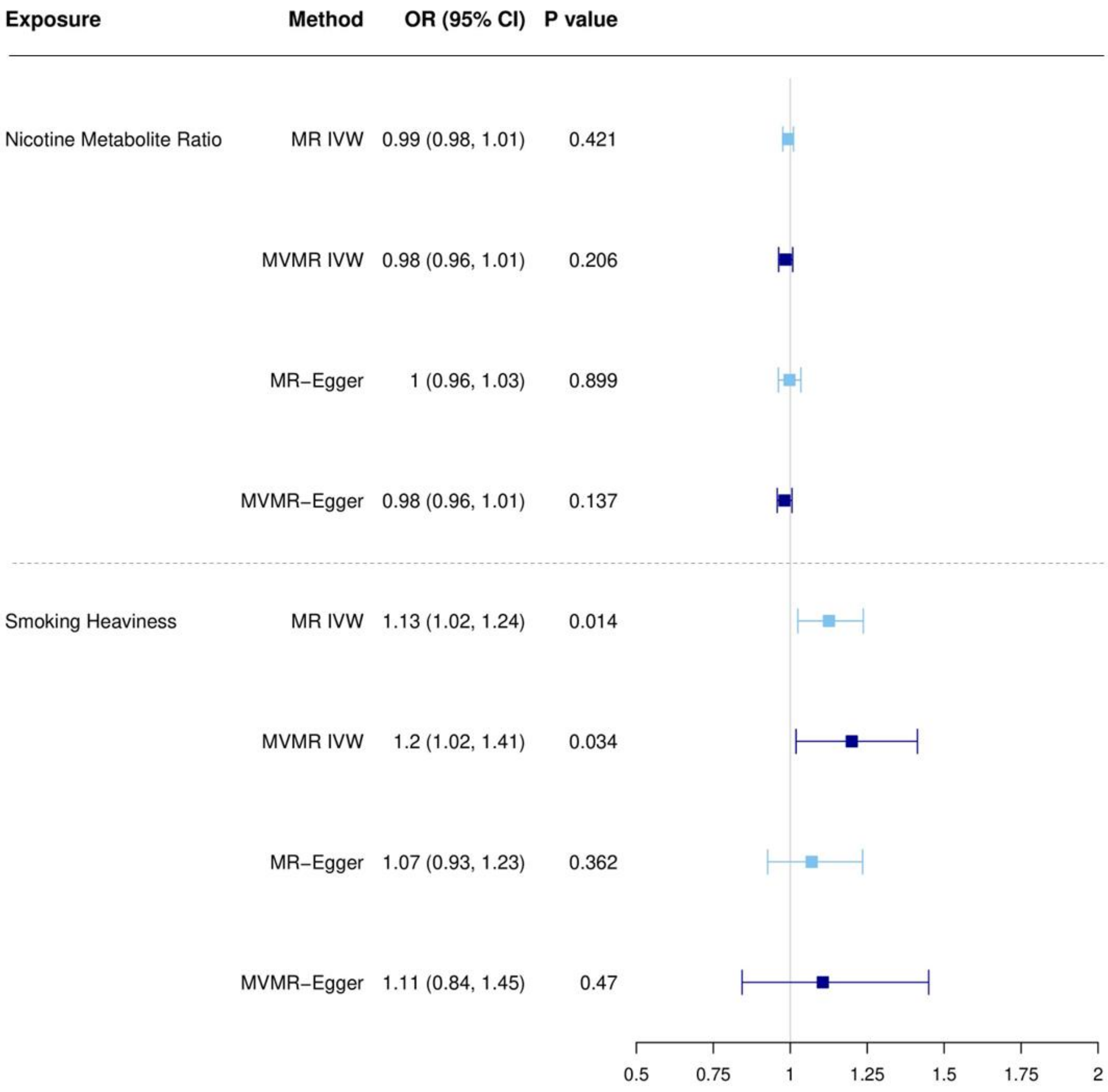

**Supplementary Figure S11.** Leave-one-out analyses depicting the results of Inverse Variance Weighted (IVW) Mendelian randomization analyses from liability to nicotine metabolite ratio on MDD risk among ever smokers after excluding each of the genetic variants from the analysis, one at the time.

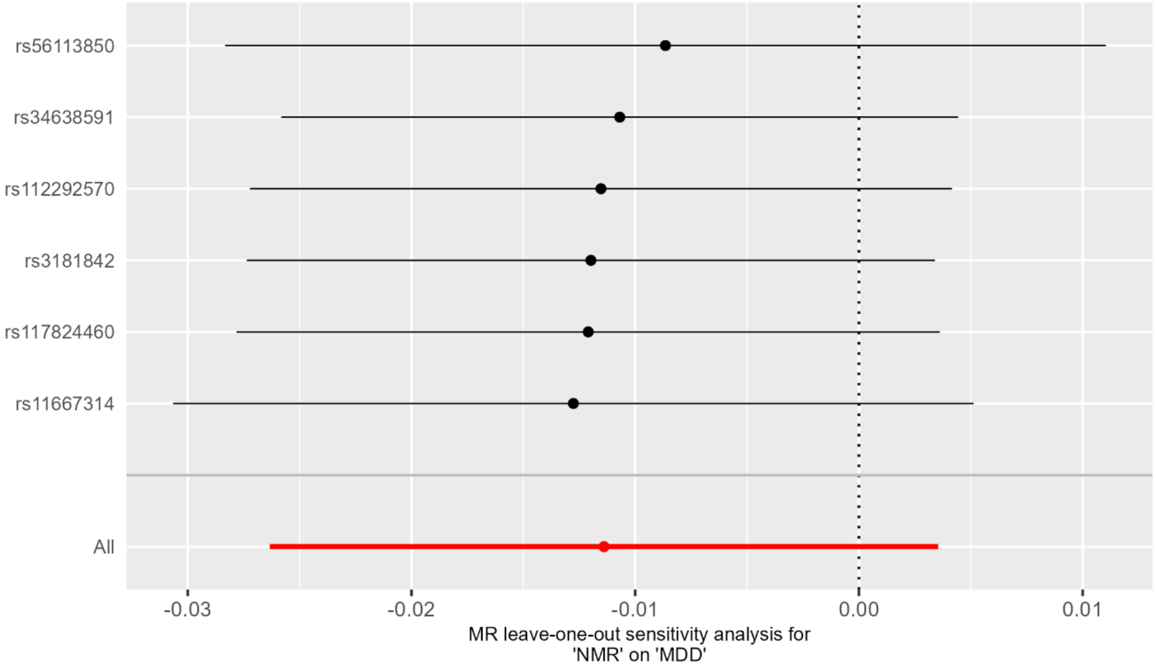

**Supplementary Figure S12**. Leave-one-out analyses depicting the results of Inverse Variance Weighted (IVW) Mendelian randomization analyses from liability to smoking heaviness on MDD risk among ever smokers after excluding each of the genetic variants from the analysis, one at the time.

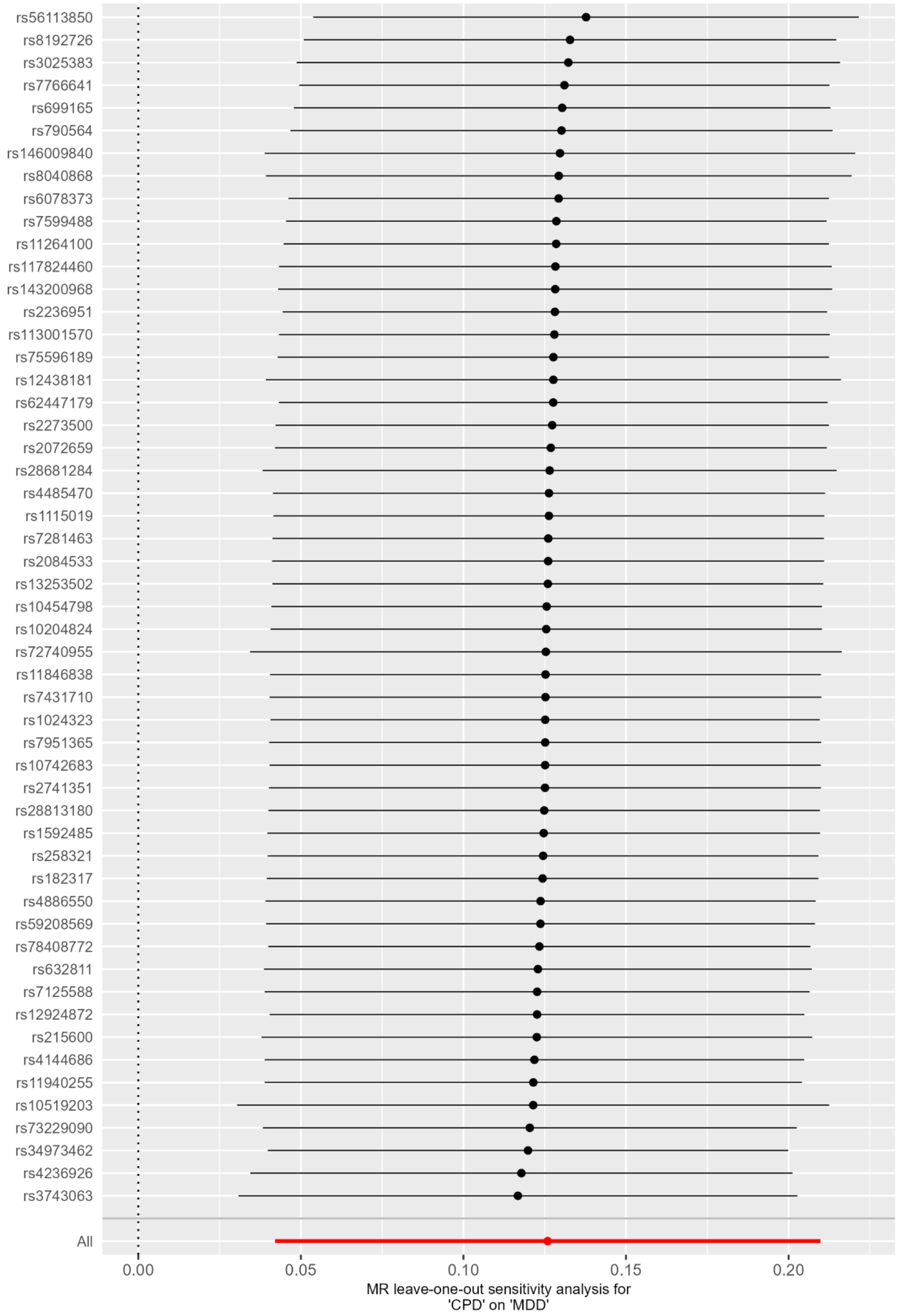
